# Contrasting genomic epidemiology between sympatric *Plasmodium falciparum* and *Plasmodium vivax* populations

**DOI:** 10.1101/2024.04.22.24306174

**Authors:** Philipp Schwabl, Flavia Camponovo, Collette Clementson, Angela M. Early, Margaret Laws, David A. Forero-Peña, Oscar Noya, María Eugenia Grillet, Mathieu Vanhove, Frank Anthony, Kashana James, Narine Singh, Horace Cox, Reza Niles-Robin, Caroline O. Buckee, Daniel E. Neafsey

## Abstract

The malaria parasites *Plasmodium falciparum* and *Plasmodium vivax* differ in key biological processes and associated clinical effects, but consequences on population-level transmission dynamics are difficult to predict. This co-endemic malaria study from Guyana details important epidemiological contrasts between the species by coupling population genomics (1,396 spatiotemporally-matched parasite genomes) with sociodemographic analysis (nationwide patient census). We describe how *P. falciparum* forms large, interrelated subpopulations that sporadically expand but generally exhibit restrained dispersal, whereby spatial distance and patient travel statistics predict parasite identity-by-descent (IBD). Case bias towards working-age adults is also strongly pronounced. *P. vivax* exhibits 46% higher average diversity (π) and 6.5x lower average IBD. It occupies a wider geographic range, without evidence for outbreak-like expansions, only microgeographic patterns of isolation-by-distance, and weaker case bias towards adults. Possible latency-relapse effects also manifest in various analyses. For example, 11.0% of patients diagnosed with *P. vivax* in Greater Georgetown report no recent travel to endemic zones, and *P. vivax* clones recur in 11/46 patients incidentally sampled twice during the study. Polyclonality rate is also 2.1x higher than in *P. falciparum,* does not trend positively with estimated incidence, and correlates uniquely to selected demographics. We discuss possible underlying mechanisms and implications for malaria control.

## Introduction

Malaria caused by the parasite *Plasmodium falciparum* is responsible for more than half a million deaths each year, predominantly in Sub-Saharan Africa and in children under five^1^. Malaria by *Plasmodium vivax* causes less acute mortality but is more widely distributed at the global scale^2,3^ and likewise causes severe morbidity and socioeconomic impact^1^.

Progress towards malaria elimination is currently stalling in many endemic regions^4^, and divergent parasite species’ responses to intervention play an important role^5–9^. *P. vivax* in particular often shows stable or increasing incidence during periods in which co-endemic *P. falciparum* is successfully being reduced^4^. It is therefore important to better understand how epidemiological differences between the species affect the impact of control strategies and whether species-tailored approaches are warranted in co-endemic settings.

*P. falciparum* and *P. vivax* are not closely related (current theory suggests 30-50 million years of independent evolution^10,11^) and differ in key developmental processes and associated clinical effects^4^. For both species, human infection begins when sporozoites enter the skin and vasculature during infected anopheline mosquito blood meal. The parasite first infects the liver and later transitions to blood-stage infection where repeated erythrocyte invasion, intra-erythrocytic replication, and erythrocyte rupture are associated with febrile and paroxysmal disease. A subset of blood-stage parasites commit to onward transmission by differentiating into the sexual, mosquito-infective gametocyte form. Meiosis occurs after transmission to the mosquito via blood meal and involves outcrossing if genetically distinct gametocytes are present (i.e., if the human infection is polyclonal). The parasite forms oocysts in the mosquito midgut and then sporozoites, which accumulate in the mosquito salivary glands^12^.

*P. vivax* behaves distinctly within the human stages of this infection cycle in multiple key aspects. First, not all *P. vivax* parasites that successfully reach the liver immediately begin schizogony, a process which begins immediately in all liver-stage *P. falciparum* parasites. Rather, a subset of liver-stage *P. vivax* parasites form a dormant hypnozoite reservoir, which can initiate additional blood-stage infections weeks to months after initial infection^13^. Clearing this relapsing hypnozoite reservoir from the liver is hindered by incomplete drug efficacy^14^, toxicity^15^, and adherence concerns^16^. In the blood stage, *P. vivax* invades a smaller subset of erythrocytes^17^ and thus more frequently creates submicroscopic infections, especially in adults^18^. *P. vivax* also develops transmissible gametocytes approximately one week earlier than *P. falciparum*. This increases the likelihood of onward transmission before antimalarial treatment occurs^19,20^. Several experimental infection studies have additionally suggested that the minimum gametocyte density required to infect susceptible mosquitoes is intrinsically lower in *P. vivax* than in P. *falciparum*^19,21–25^.

Taken together, the above properties are thought to make P. *vivax* more resilient to clinical interventions targeting symptomatic pediatric infection, a main focus of *P. falciparum* control. Vector control measures are also thought to have weaker or delayed effects on *P. vivax* due to post-intervention relapse from undetected or unsuccessfully cleared hypnozoite reservoirs, and higher probability to achieve onward transmission when mosquito abundance is low. Furthermore, long-lasting hypnozoite reservoirs and more transmissible asymptomatic blood-stage infections may make *P. vivax* more effective at long-distance dispersal and outcrossing with unrelated strains. This may also include greater potential for allochthonous transmission where malaria was previously absent or cleared^26–28^.

The epidemiological effects of these biological differences between *P. falciparum* and *P. vivax* have been difficult to quantify because synchronized comparative study designs that avoid confounding by methodological and spatiotemporal factors are challenging to establish^29^. Study systems representing true species sympatry are also rare, as *P. falciparum* and *P. vivax* often occupy distinct, only partially overlapping socio-geographic partitions within areas more broadly considered co-endemic^2,3^.

This study uniquely couples a spatiotemporally matched genomic sampling scheme with epidemiological analysis of disaggregated (patient-level) malaria case records to discern co-endemic *P. falciparum* and *P. vivax* transmission dynamics. We focus on Guyana, where both species contribute substantially to the national case count (32.3% *P. falciparum*, 67.4% *P. vivax*, and <1% *P. malariae* in 2019) and where true sympatry is geographically widespread. Malaria in Guyana also involves a diverse mix of ethnicities (e.g., strong representation from Afroguyanese, East Indian, and Amerindian ethnicities) and dynamic population mobility patterns relating to the mining field. Next to important distinctions relating to geographic and host demographic variables, we demonstrate clear species contrasts in strain ancestry, persistence, dispersal, and infection complexity within this multifaceted transmission context. We discuss possible biological drivers behind divergent species epidemiologies and identify various implications for intervention.

## Results

### General spatiotemporal species prevalence

We first conducted a descriptive summary of country-wide passive case detection records made available by the Guyana Ministry of Health (GMOH) for 2006 to 2019. Rolling monthly counts demonstrate well-balanced case burden between species from 2006 to 2010 followed by higher *P. falciparum* incidence from 2010 to 2013 (Fig. 1a). *P. vivax* cases began to exceed those of *P. falciparum* in mid-2013 and remained overrepresented until 2019. Individual-level case metadata have been curated by the GMOH for 2019. We therefore focus on 2019 for most subsequent epidemiological analyses. 2019 records include 17,710 single-species cases (67.4% *P. vivax*, 32.3% *P. falciparum*, and 0.2% *P. malariae*) and 1,001 mixed-species cases containing both *P. vivax* and *P. falciparum* (Supplementary Table 1). Both species show transmission throughout the year and elevated incidence between April and August (Supplementary Fig. 1).

**Fig. 1.**
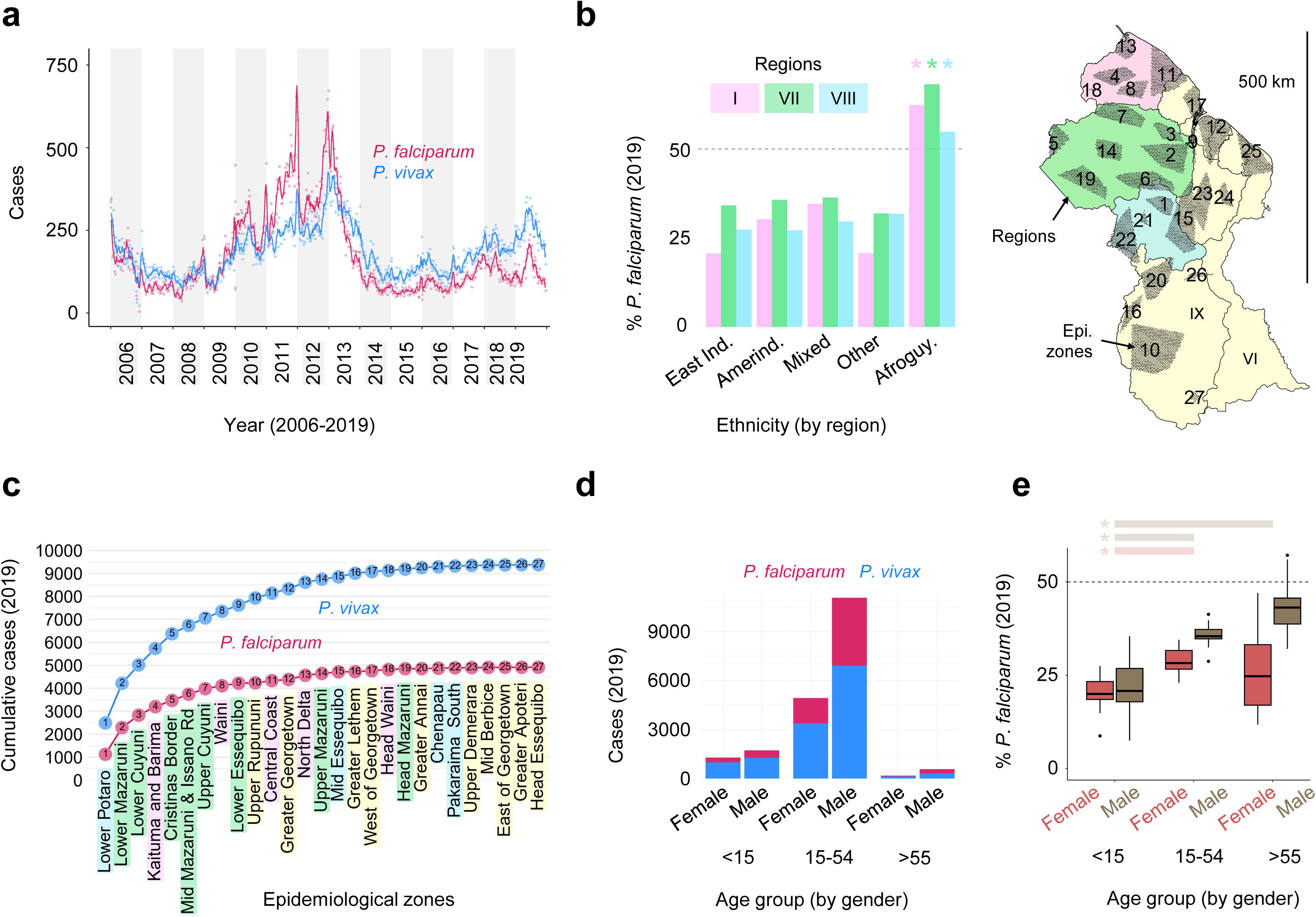
Spatiotemporal and demographic patterns of *P. falciparum* and *P. vivax* in Guyana. a) Weekly reported cases (points) and 30-days rolling average (line) between 2006 and 2019. **b)** Percent of cases representing *P. falciparum* (% Pf) by region and ethnicity in 2019. Asterisks indicate significant differences in % Pf between Afroguyanese and non-Afroguyanese cases in each region (χ^2^-test). **c)** Cumulative case count in epidemiological zones with >500 total cases in 2019. Zones on the y-axis are ordered by *P. vivax* case count (descending from top to bottom). **d)** Case counts by gender and age in 2019. **e)** % Pf by gender and age in 2019. Asterisks indicate significant differences between age groups (Dunn test).

### Species intricacies of a wide co-endemic distribution

2019 case records show the extent of *P. falciparum* – *P. vivax* geographic and demographic co-endemicity in Guyana (Fig. 1b-d). Regions I, VII, and VIII account for 94.6% of *P. falciparum* cases and 89.3% of *P. vivax* cases, and regional species proportions remain stable across East Indian, Amerindian and Mixed/Other ethnicity groups. While *P. vivax* consistently predominates in these groups, case majority flips to *P. falciparum* in the Afroguyanese (Chi-squared tests, χ^2^ = 36.13 (I), 269.63 (VII), 125.51 (VIII), p < 0.001 (all regions), Fig. 1b). We further classified infection localities into 27 epidemiological zones (average 3,631 km^2^ each) defined using river-road transport networks and mobility friction maps (Fig. 1c, Supplementary Fig. 2). Again, species sympatry is consistently maintained in all zones within Regions I, VII, and VIII. The relative proportion of *P. vivax* cases is generally higher outside of this main malaria range, especially to the south in Region IX (e.g., Upper Rupununi and Greater Lethem). Patients aged 15-54 clearly predominate in both species but are more common in *P. falciparum* (84.5%) than in *P. vivax* (79.2% – Chi-squared test, χ^2^ = 82.89, p < 0.001, Fig. 1d). The proportion of cases representing *P. falciparum* also increases markedly with age in male patients (Kruskal-Wallis test, H = 23.64, p < 0.001 – see post-hoc significance in Fig. 1e). Working-age (≥15 years) to child (<15 years) case ratios are less markedly elevated in Amerindians than in non-Amerindian ethnicities (3.8 to 1 vs. 13.5 to 1, respectively, for *P. falciparum* (Chi-squared test, χ^2^ = 273.84, p < 0.001) and 2.6 to 1 vs. 7.4 to 1, respectively, for *P. vivax* (Chi-squared test, χ^2^ = 515.98, p < 0.001), Supplementary Fig. 4). Elevated case bias towards non-Amerindian working-age males is consistent with gold mining driving malaria burden in Guyana^30^, especially in non-resident (e.g., ‘coast lander’^31^) mining groups. Higher working-age to child case ratio for *P. falciparum* vs. *P. vivax* may also suggest that transmission from mining to non-mining communities, and sustained transmission in non-mining communities, is less common in *P. falciparum* than in *P. vivax*.

### Deep genomic profiling of each species across 2020 and 2021

To search for epidemiological features that are not observable from traditional symptomatic case analysis, we sequenced 708 *P. falciparum* and 762 *P. vivax* genomes from patient blood spots collected passively by the GMOH. Collection followed informed consent using a study protocol approved by ethical committees of Harvard University and the government of Guyana. Sequencing was successful for 666 *P. falciparum* and 705 *P. vivax* genomes (Supplementary Table 2) continuously spanning January 2020 to June 2021 (Fig. 2). Of these 1,371 samples, 1,216 (89%) are coupled with patient travel history records used to infer geographic infection source (representing Regions I, VII, and VIII in 1,156 (95.1%) samples). We additionally sequenced 7 *P. falciparum* and 16 *P. vivax* samples from 2019 representing infections from Venezuela and analyzed two publicly available Venezuelan *P. falciparum* genomes from 2015/16 (European Nucleotide Archive (https://www.ebi.ac.uk) accessions ERR1818176 and ERR2496572).

**Fig. 2.**
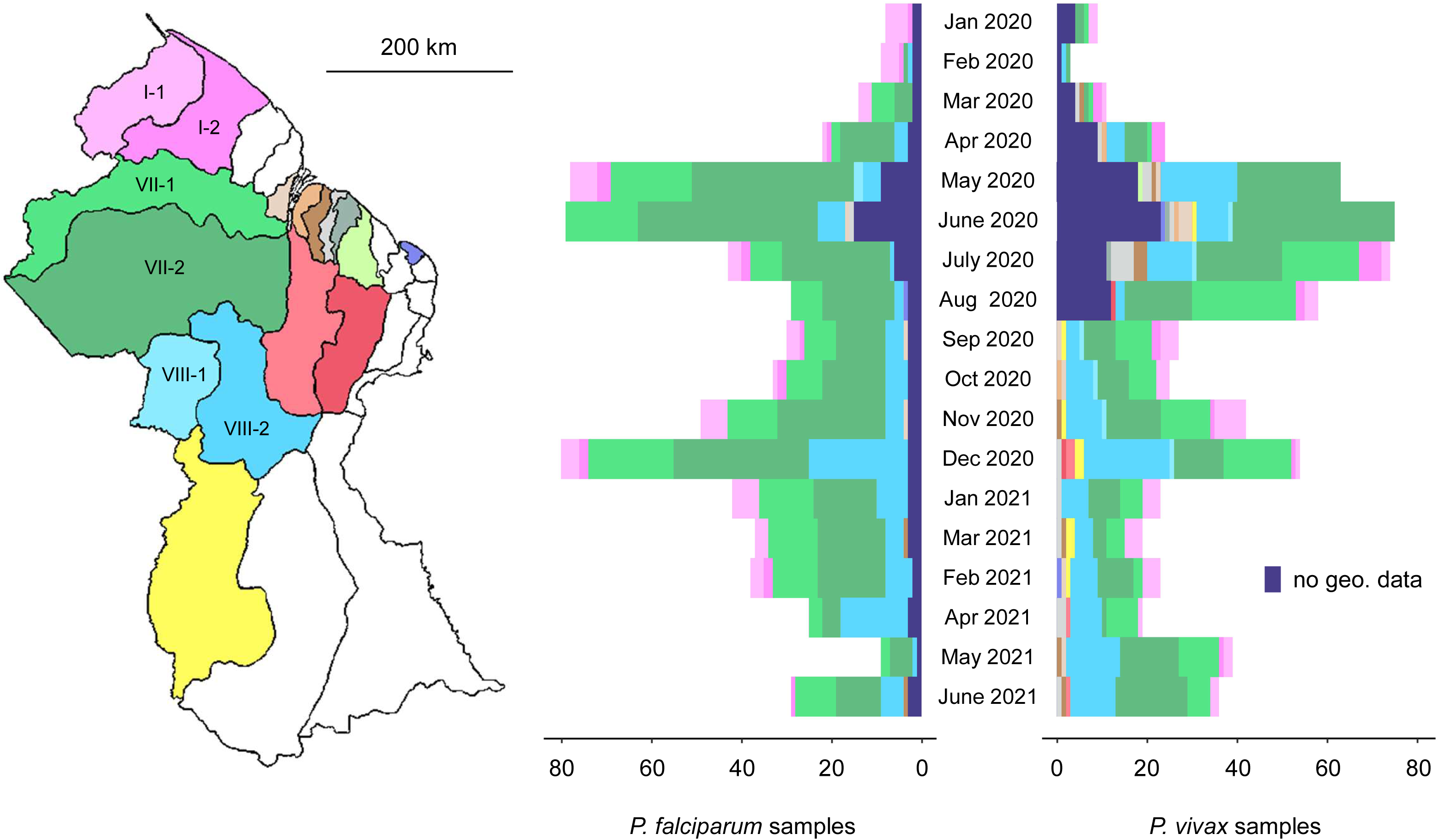
Spatiotemporally matched *P. falciparum* and *P. vivax* sampling in Guyana. Colors in the map and histograms represent the neighborhood councils (NDCs) to which infections from 2020-21 were attributed based on patient response about location of stay 14 days prior to diagnosis. NDCs within Regions I, VII, and VIII are labeled in the map.

### Discrepant genomic diversity and relatedness between species

Genomic analyses focused on 46,067 and 266,141 sites with segregating single-nucleotide polymorphisms (SNPs) identified in the *P. falciparum* and *P. vivax* sample sets (respectively) following quality filtration steps (see Methods). This difference in the number of segregating sites and in average pairwise nucleotide diversity observed within each sample set (1.99 ⋅ 10^-4^ differences per bp in *P. falciparum* and 2.91 ⋅ 10^-4^ differences per bp in *P. vivax,* Supplementary Fig. 5) align with previous evidence that genetic diversity is higher in *P. vivax* at the global scale^32^.

To further characterize diversity within each sample set, we applied a hidden state model^33^ to estimate parasite relatedness due to recent common ancestry (identity-by-descent; IBD) for all monoclonal sample pairs (we excluded 250 samples identified as multi-strain infections using a Bayesian framework^34^). The relatedness distributions (Fig. 3a) clearly differ between the two species (Kolmogorov-Smirnov test, D = 0.99, p < 0.001). Pairwise relatedness values in *P. vivax* exhibit a mean of 0.048, with very little standard deviation (sd = 0.031) and minimal skew or outlier occurrence. These results are indicative of a diverse, frequently outcrossing population without substantial sub-clustering into groups of highly related individuals. In contrast, most pairwise comparisons in *P. falciparum* exhibit greater than half-sibling-level relatedness (mean = 0.310 ± 0.124 sd). The broader, right-tailed relatedness distribution also includes a peak near 1 representing the presence of identical clones as well as values between 0.5 and 0.75 representing the presence of various inbred sibling-level relationships. A network analysis identified 79 clonal groups (>0.90 IBD) among *P. falciparum* samples, represented by 2 to 39 members each (Supplementary Fig. 6). Clonal group membership detection was often spatiotemporally aggregated (e.g., >50% of group members detected within 4 months per epidemiological zone, Supplementary Fig. 7), consistent with outbreak-like patterns of prevalence^35^. The largest clonal groups spread across multiple geographic areas (Supplementary Fig. 8a), and most appear highly interrelated (e.g., 50 groups collapse into a single network when re-clustering samples at >0.60 IBD, Supplementary Fig. 6). Four groups noted with asterisks in Supplementary Fig. 6 and mapped in Supplementary Fig. 8b appear slightly divergent (1.7 to 3.3 sd below average between-group IBD). Other clonal patterns mapped in Supplementary Fig. 8b include the clustering of groups #7, #9, and #10 around Port Kaituma and Mabaruma (Region I), groups #18, #24, and #49 along the Mazaruni and Puruni Rivers (Region VII), and groups #27, #32, and #52 around Mahdia and the lower Potaro River (Region 8). Two divergent clonal pairs (#20 and #65) also have partial membership in Venezuela (star symbols in Supplementary Fig. 8b) and likely represent transmission of imported parasite lineages (<u>Supplementary Text 1</u>, Supplementary Fig. 9). Group #20 also represents a case of longer-term clonal persistence from 2015 to 2020.

**Fig. 3.**
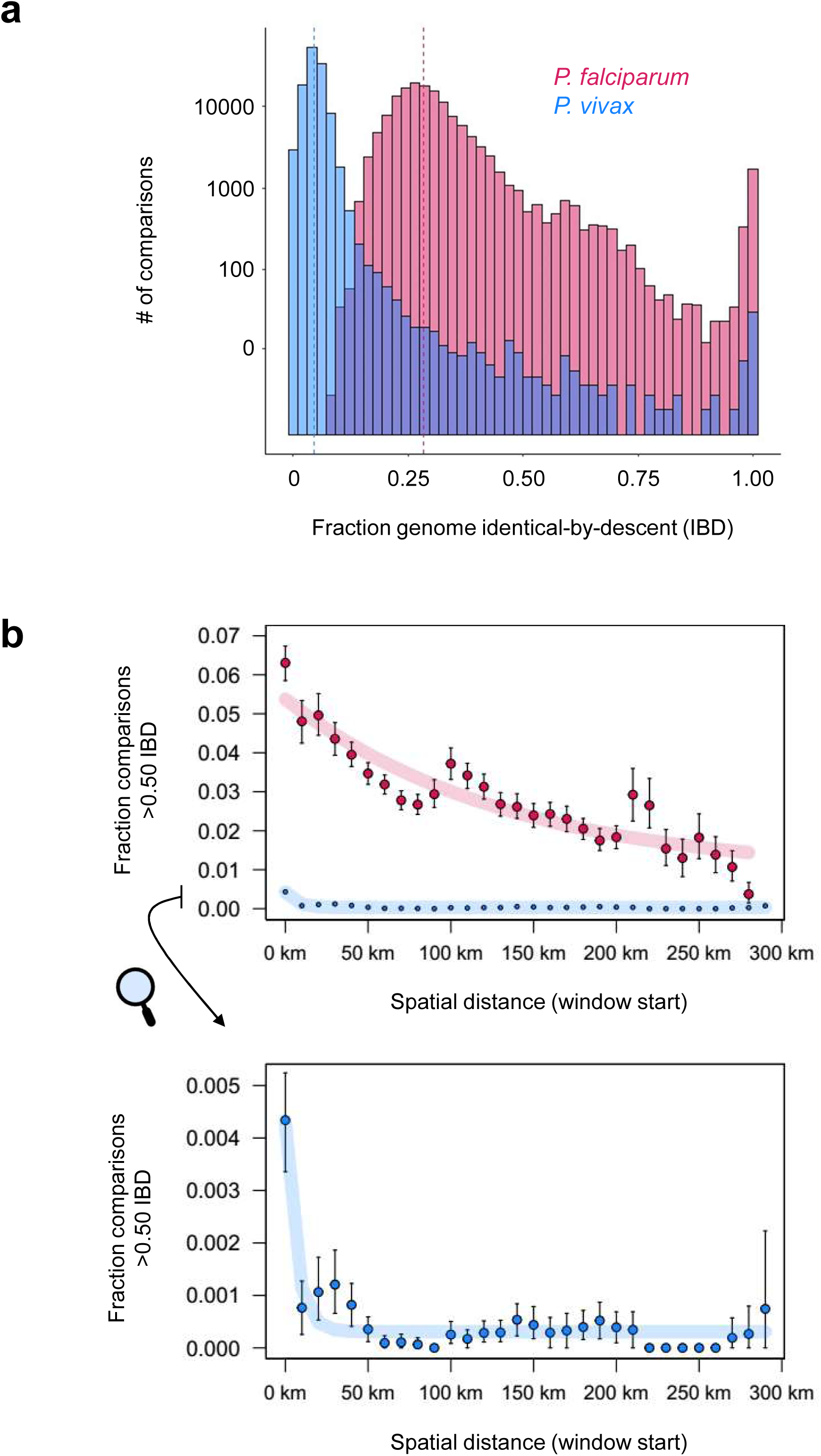
Pairwise IBD and relationships to spatial distance in *P. falciparum* and *P. vivax* in Guyana. a) Histogram of pairwise IBD in 2020-21. The y-axis is log10-scaled to help display less frequent close-relative observations. Dashed lines indicate species medians. **b)** Relationship of spatial distance between inferred infection sites and the frequency of >0.50 IBD in 2020-21. Windowed calculation using step size = 10 km and starting at 0-30 km. Error bars represent 90% confidence intervals from bootstrapping 1000x. The model y = a * e^(-b * x) + c was used to fit regression lines of linear and logarithmic shape. Clonal *P. falciparum* groups represented by ≥10 members are excluded from distance analyses.

Clonal network analysis results for *P. vivax* are very different. We observe only 29 clonal groups, of which 23 contain 2 members each and 6 contain 3 members each. Post-hoc review of patient-specific case codes (blinded to non-GMOH authors) also clarifies that 11 of 23 clonal pairs represent repeated sampling from the same patient (median time between 1st and 2nd visit = 109 days (minimum = 40 days, maximum = 229 days). These likely represent relapses registered as new infections prior to genetic analysis. No close relatedness occurs among any clonal groups (the same number of groups created via clustering at >0.90 IBD remains when re-clustering at >0.30 IBD) or between Guyanese and Venezuelan sample sets (Supplementary Fig. 6, <u>Supplementary Text 1</u>, Supplementary Fig. 9). Furthermore, clonal persistence in *P. vivax* (average = 79 days, maximum = 243 days) appears more temporally limited (Wilcoxon test, W = 55078, p = 0.003) than in *P. falciparum* (median = 113.5 days, maximum = 500 days, excluding imported lineages (groups #20 and #65), Supplementary Fig. 10).

### Regional isolation-by-distance in P. falciparum vs. micrographic structure in P. vivax

We next evaluated whether parasite genetic relatedness declines with spatial distance (Fig. 3b). The presence of ‘isolation-by-distance’ is a common indicator of population structure due to spatially limited dispersal, understanding of which is key to the success of control objectives such as limiting the spread of drug resistance^36,37^. We applied sliding windows of spatial distance, quantifying the fraction of pairwise comparisons in each 30 km window that showed >0.50 IBD. Results for *P. falciparum* demonstrate a linear decrease in the frequency of >0.50 IBD as spatial distance increases between sample pairs (Pearson’s r = 0.89, p < 0.001). Exceptions occur at window starts near 100 km and 200 km where the frequency of close relatives briefly rebounds to levels observed at shorter distance classes. These upticks may reflect accumulation of relatedness between common travel hubs or mining foci and discontinuous transmission risk during interregional host movement.

In *P. vivax*, sample pairs with >0.50 IBD were very rare (n = 87) but also clearly overrepresented in the first spatial distance class (0-30 km, Fig. 3). A continued decline in >0.50 IBD frequency with spatial distance was however not apparent. To help assess whether the spatial distance across which negative correlation between relatedness and spatial distance persists differs between species, we quantified 99^th^ percentile (p99) IBD, an alternative definition of close ancestry that normalizes statistical power in comparative Mantel tests (Supplementary Fig. 11, top). Correlogram results for *P. falciparum* indicated significant correlation between genetic distance (1 - p99 IBD frequency) and spatial distance for 0-30 km (Mantel r = 0.49, p = 0.001) as well as for 30-60 km distance classes (Mantel r = 0.19, p = 0.017). In *P. vivax*, correlation between 1 - p99 IBD frequency and spatial distance loses statistical significance at 30-60 km (Mantel r = 0.12, p = 0.089). Shorter isolation-by-distance signal relative to *P. falciparum* also occurred when using the absolute, >0.50 IBD metric to define close ancestry (Supplementary Fig. 11, bottom).

### Genetic vs. non-genetic markers accord better on P. falciparum than on P. vivax dispersal

We further examined how parasite pairs exhibiting >0.50 IBD were distributed among epidemiological zones (Fig. 4) and whether this parasite genetic structure correlated to patient travel patterns inferred by comparing sites of patient diagnosis and prior stay (2 weeks) in the epidemiological database from 2019. Examining the extent to which parasite propagation across specific geographic or administrative units can be tracked and predicted using genetic or traditional case metrics is important in helping determine potential to focus and tailor intervention activities and surveillance data types.

**Fig. 4.**
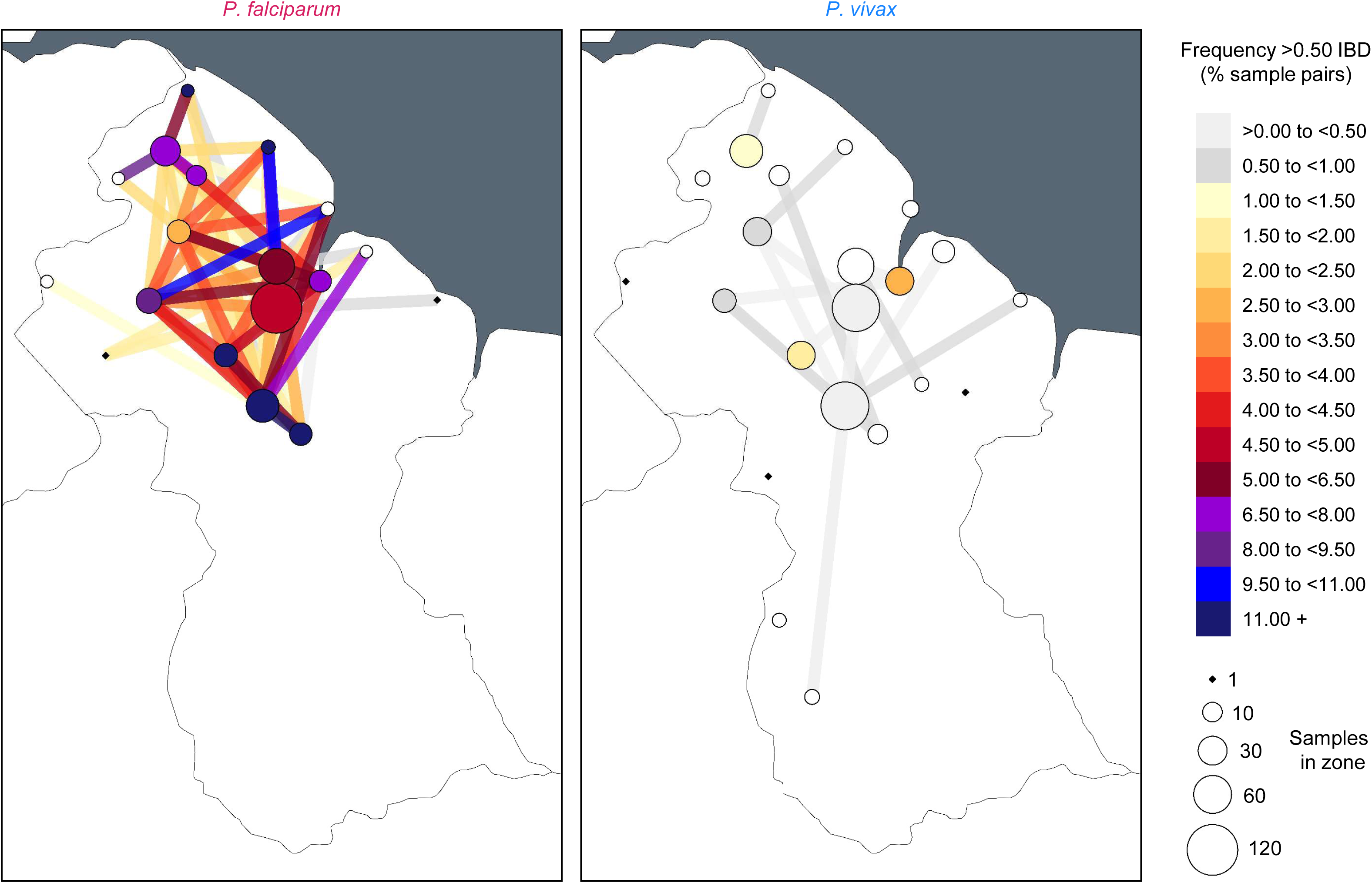
Frequency of >0.50 IBD within and between epidemiological zones in *P. falciparum* and *P. vivax* in Guyana. Nodes indicate epidemiological zones. Node and segment colors indicate frequency of >0.50 IBD within and between epidemiological zones (respectively) in 2020-21. Node sizes are proportional to genomic sampling size. Between-zone comparisons represented by ≤50 comparisons are excluded.

Corollary to the clear elevation in >0.50 IBD frequency at 0-30 km (Fig. 3) and significant isolation-by-distance at 0-30 km (Supplementary Fig. 11) in both *P. falciparum* and *P. vivax*, both also generally showed highest frequencies of elevated relatedness within (i.e., not between) epidemiological zones (Fig. 4). This observation from the genomic data matches the observation from the epidemiological database that the majority of symptomatic malaria cases involve non-mobile, ‘local’ infections (black pie slices, Supplementary Fig. 12), i.e., patients reporting absence of travel 14 days prior to diagnosis. The epidemiological data also suggest that the proportion of local to nonlocal infections (i.e, cases in which patients reported having stayed in a distinct epidemiological zone 14 days prior to diagnosis) is higher for *P. vivax* than for *P. falciparum* in 16 of 23 analyzed epidemiological zones (Supplementary Fig. 13). These 16 zones include non-endemic coastal areas such as Greater Georgetown, where 178 of 1,611 (11.0%) *P. vivax* patients (vs. 45 of 867 (5.2%) *P. falciparum* patients) reported absence of prior travel (Chi-squared test, χ^2^ = 22.915, p < 0.001). Elevated pairwise nucleotide diversity (+20.7% π) in *P. vivax* samples attributed to Greater Georgetown (Welch’s t test, t = 4.71, p < 0.001, Supplementary Fig. 5) further supports the hypothesis that relapses representing various different geographic infection sources are diagnosed in this capital district.

To statistically compare inference of parasite movement based on genetic vs. epidemiological data types, we correlated the relative frequency of parasite >0.50 IBD and patient travel events (‘case flow’) among all pairs of endemic epidemiological zones for which patient movement was detected in the epidemiological database (i.e., all arrowed segments in Supplementary Fig. 12). In the case of *P. falciparum*, relative frequencies of >0.50 IBD and case flow were moderately correlated (Pearson’s r = 0.44, p = 0.029) between endemic zones (Fig. 5), with most conspicuous congruence in the context of reduced connectivity between Region I and Regions VII and VIII (e.g., see short spokes of elevated >0.50 IBD frequency and intensified case flow around Kaituma and Barima in Region I, yet limited connectivity farther South in Fig. 4 and Supplementary Fig. 12). Only 0.8% of nonlocal infections sampled in Regions VII and VIII (grey pie slices, Supplementary Fig. 12) represented infection sites from Region I. Case flow to Greater Georgetown was also weaker for infections attributed to Region I (36.4%) than for infections attributed to Region VII (43.4% – Chi-squared test, χ^2^ = 48.22, p < 0.001) or to Region VIII (74.1% – Chi-squared test, χ^2^ = 874.35, p < 0.001). These results may in part reflect the absence of efficient travel routes from Region I to Georgetown (East) and to Regions farther South. Rivers key to travel in Region I (e.g., the Barima and Barama) lead into a Northwest delta while those key to travel in Regions VII and VIII (e.g., the Cuyuni and Mazaruni) flow East into the Essequibo near Georgetown. A handful of semi-developed roads also parallel and connect areas of the Cuyuni, Mazaruni, and Essequibo in Regions VII and VIII.

**Fig. 5.**
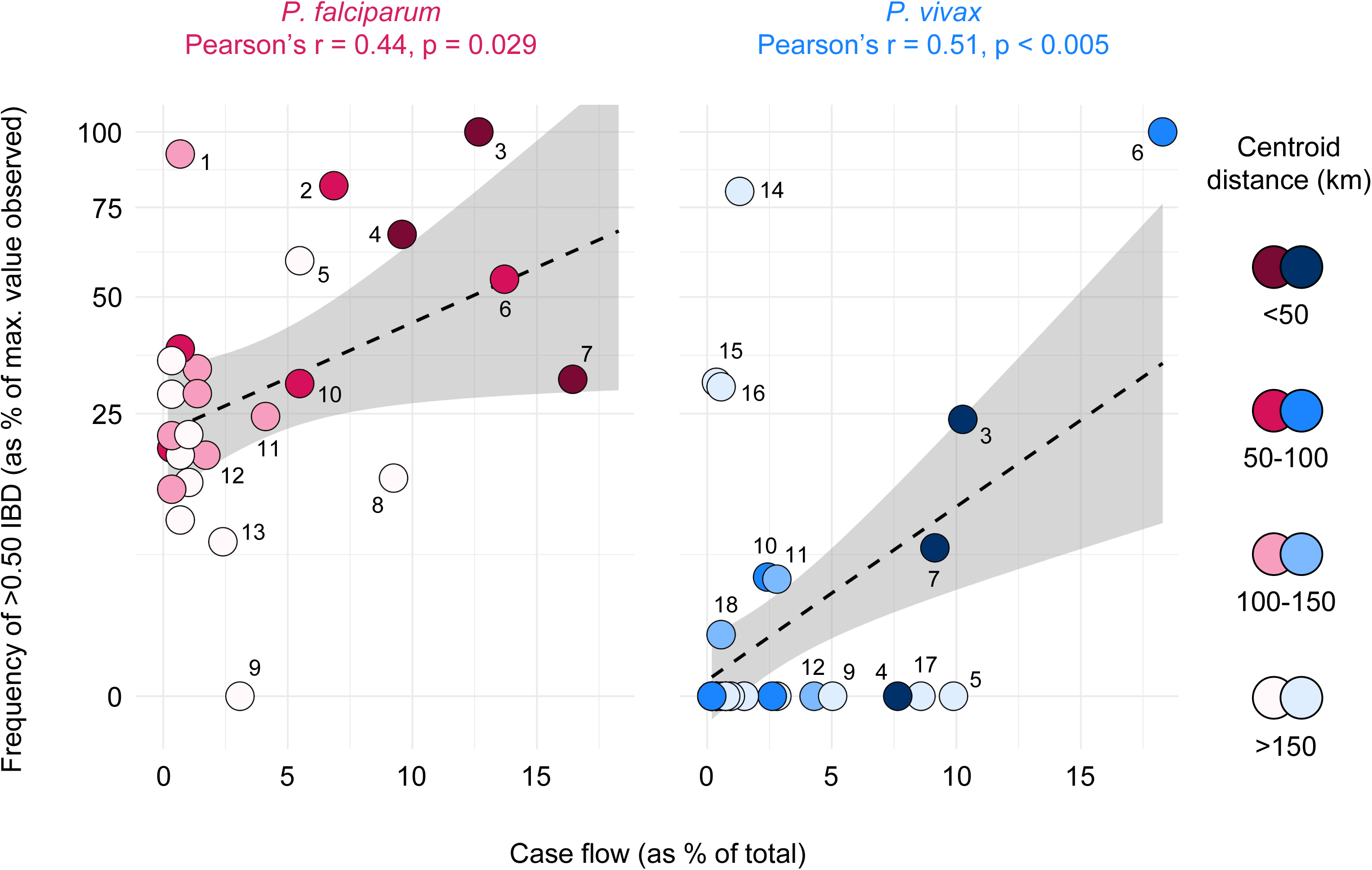
Relationship between patient case flow and the relative frequency of >0.50 IBD in *P. falciparum* and *P. vivax* in Guyana. Genetic data (y-axis) represents 2020-21 and epidemiological (case flow) data (x-axis) represents 2019. Point color indicates the spatial distance separating the zones being compared. Comparisons represented by ≤50 comparisons excluded. Grey shading indicates 95% confidence intervals predicted by linear regression (dashed line). Labeled points represent 1) Central Coast (CC) to Lower Cuyuni (LC), 2) Head Waini (HW) to Kaituma and Barima (KB), 3) Lower Potaro (LP) to Mid Essequibo, 4) KB to Waini, 5) CC to Lower Mazaruni (LM), 6) KB to North Delta (ND), 7) LC to LM, 8) Head Mazaruni to LM, 9) Cristinas Border to LM, 10) LM to Mid Mazaruni and Issano Rd, 11) LM to Upper Cuyuni, 12) CC to KB, 13) LM to ND, 14) East of Georgetown to LP, 15) LM to Upper Rupununi (UR), 16) LP to UR, 17) Greater Lethem to LM, and 18) LM to LP.

For *P. vivax*, the frequency of >0.50 IBD also correlated significantly with case flow between endemic zones (Pearson’s r = 0.51, p < 0.001, Fig. 5). Unlike in *P. falciparum*, however, significance is lost when broadening the classification of elevated relatedness to p99 IBD (Supplementary Fig. 14). This observation helps clarify that the significant relationships detected when regressing >0.50 IBD frequency on spatial distance (Fig. 3) and case flow (Fig. 5) do not mean that the *P. vivax* population is predictably structured as a whole. Results are instead consistent with a dispersive, frequently outcrossing *P. vivax* population in which pairwise relatedness is swiftly erased and local genotype associations are rarely established. In this scenario, the ephemeral presence of intact clones or sibling-level relatives creates micrographic spatiotemporal autocorrelations that are not representative of broader population dynamics.

### Higher outcrossing potential in P. vivax – do relapse and risk carryover play a role?

We further assessed indications from relatedness analyses that outcrossing rate is elevated in *P. vivax* by examining polyclonality rate (i.e., the fraction of samples containing multiple distinct strains) and its variation with respect to geographic and demographic variables. Polyclonality rate is closely related to outcrossing rate because parasite sexual recombination in the mosquito after blood meal only results in outcrossing if multiple distinct strains are ingested simultaneously from the human host.

We observed a polyclonality rate of 24.0% in *P. vivax* vs. 11.4% in *P. falciparum* (Chi-squared test, χ^2^ = 36.72, p < 0.001, Fig. 6a). Elevated *P. vivax* outcrossing rate implied by this 2.1x polyclonality rate differential helps explain the aforementioned paucity in clonal relationships (Fig. 3) and their limited persistence over time (Supplementary Fig. 10). It is also consistent with the observation that linkage disequilibrium between SNPs declines across physical genetic distance more readily in *P. vivax* than in *P. falciparum* (Supplementary Fig. 15).

**Fig. 6.**
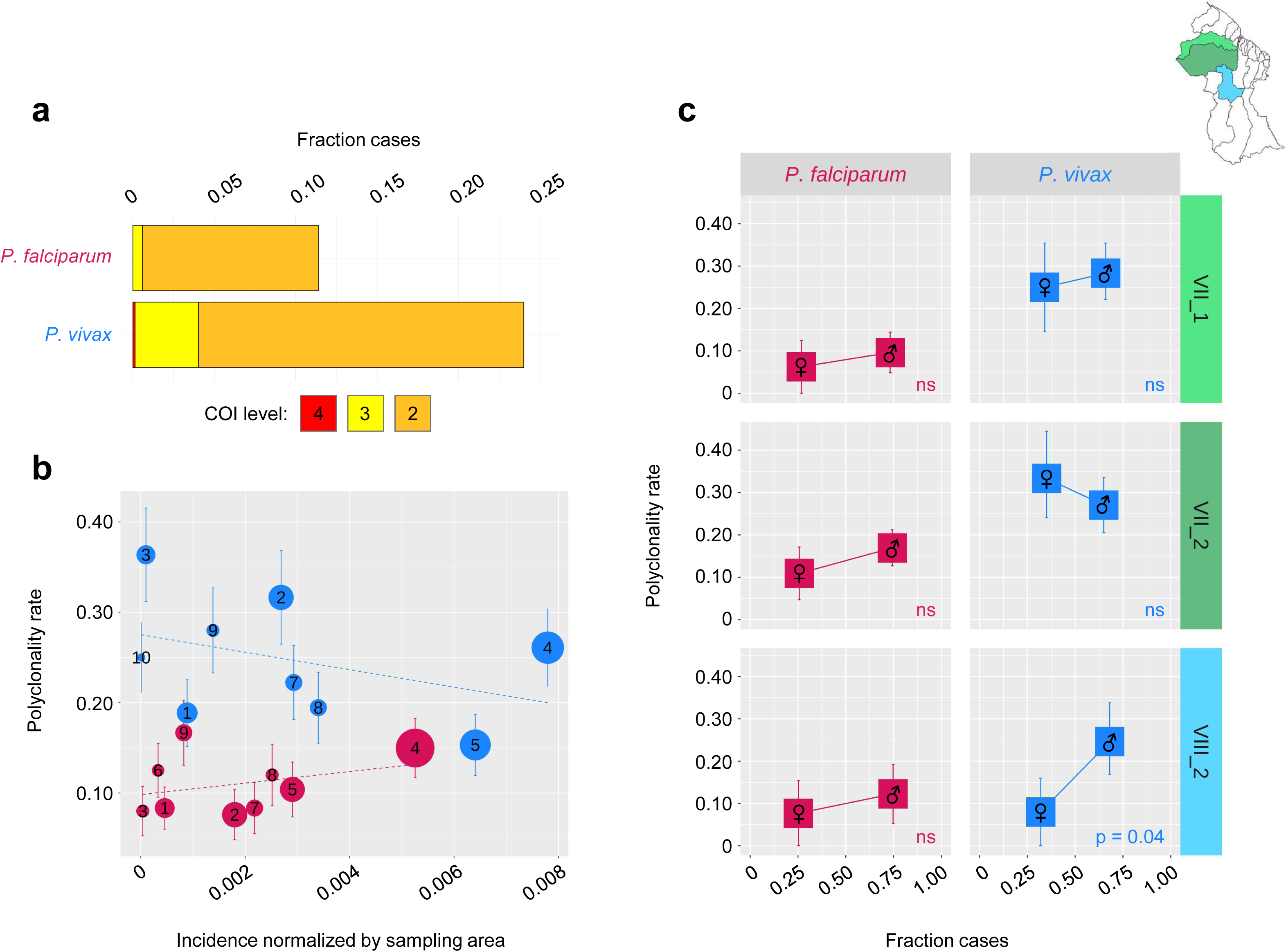
Polyclonality rates and relationships to estimated incidence and patient gender in *P. falciparum* and *P. vivax* in Guyana. a) Complexity of infection (COI) values in 2020-21, population-wide. **b)** Polyclonality rate (2020-21) vs. estimated incidence (2019) in epidemiological zones represented by ≥ 20 genomic samples (Kaituma_and_Barima (1), Lower_Cuyuni (2), Lower_Essequibo (3), Lower_Mazaruni (4), Lower_Potaro (5), Mid_Essequibo (6), Mid_Mazaruni_and_Issano_Rd (7), Upper_Cuyuni (8), Upper_Mazaruni (9), Greater_GT (10)). Error bars represent 95% confidence intervals from bootstrapping 100x. **c)** Polyclonality rate (2020-21) vs. patient gender in neighborhood district councils (NDCs) represented by ≥20 genomic samples. Error bars represent 90% confidence intervals from bootstrapping 1000x.

Polyclonality rate is considered a positive correlate of *P. falciparum* transmission intensity in the absence of high rates of case importation^38^. We assessed this possible relationship in *P. falciparum* and *P. vivax* (in which polyclonality may additionally occur via hypnozoite-based relapse) by plotting polyclonality rates per epidemiological zone against corresponding incidence estimates (Fig. 6b, see Methods). Interestingly, *P. vivax* polyclonality rates remain consistently inflated, regardless of incidence estimate. Polyclonality rate and incidence estimates also trend in opposite directions for *P. falciparum* vs. *P. vivax*, though neither achieves statistical significance. Polyclonality rate was highest in *P. vivax* infections from the Lower Essequibo zone, a key area of travel convergence from the hinterlands of Region VII but one in which malaria risk is not considered high. These results may indicate that *P. vivax* samples from areas of lower endemicity are more likely to represent imported cases (possibly in the form of relapse) that involve multiple accumulated strains.

To further understand polyclonality rate relationships to transmission risk, we compared polyclonality rates in ≥15 year-old male and females within the three neighborhood councils (NDCs) containing ≥20 genomic samples for each of these groups (Fig. 6c). Males represented the majority of epidemiological case records in the three NDCs, suggesting that transmission risk is generally higher in male-associated occupations. Polyclonality rate in *P. falciparum* thus followed expectations in that observed values appeared consistently higher in males than in females. Polyclonality rates in *P. vivax* were generally more variable (see confidence intervals, especially in females (Fig. 6c)), and were higher in females than in males in NDC VII-2, an area that contains Mahdia and various popular mining sites (e.g., the Mazaruni River Quarry and the Karouni Project). Interestingly, this trend was driven by polyclonality rate enrichment in females among patients reporting Venezuelan nationality (Chi-squared test, χ^2^ = 4.33, p = 0.037, Supplementary Fig. 16), possibly because infection risk is enhanced in female-biased occupations that accompany the active mining sector (e.g., in rest/supply areas where frequent contact among gametocyte-carrying male miners, mosquitoes, and female non-miners occurs). Enhanced *P. vivax* transmissibility, e.g., due to presymptomatic gametocytemia, may heighten this risk.

## Discussion

In this study we coupled comparative malaria parasite population genomics with passive case record metadata analyses to compare co-endemic *P. falciparum* and *P. vivax* epidemiology in Guyana. We observed several key genomic epidemiological contrasts that indicate the need to consider species-specific approaches to malaria intervention.

Most notably, we observed dramatically different patterns of genomic relatedness between the two species. The *P. falciparum* sample set exhibited a relatively broad pairwise IBD distribution centered near values expected for half-sibling-level relatedness, whereas in *P. vivax* this distribution was less variable and clearly shifted towards zero (6.4x lower average IBD). Clonal pairwise relationships (>0.90 IBD) were detected >50x more often in *P. falciparum* and most clonal *P. falciparum* groups exceeded 3 samples in membership size (max. 38). Many of these groups also displayed spatiotemporally aggregated, outbreak-like patterns of detection. In *P. vivax*, clonal groups had no more than 2-3 members and appeared to persist for shorter periods of time; many clonal *P. falciparum* relationships were detected between samples separated by >12 months whereas for *P. vivax* the maximum time between clone detection was 243 days.

This strong contrast in relatedness structure may derive mechanistically from polyclonality rate differences between the species. Polyclonality rate is a key determinant of the extent to which clones and near-relatives can persist in transmission cycles because polyclonality is a prerequisite to sexual recombination between distinct strains (outcrossing) in the mosquito stage. In this study, *P. vivax* polyclonality was not only 2.1x higher than in *P. falciparum* at the population level, but also remained consistently inflated when accounting for estimated incidence in subgeographic comparisons. This result suggests that elevated polyclonality rates in *P. vivax* are not a product of higher infection incidence alone. *P. vivax* relapse from the dormant hyponozite liver stage likely represents an additional source of polyclonality because relapse can overlap with current infection and can involve the successive activation of divergent strains^39^.

Importantly, though humans are the common agent of dispersal throughout Guyana for both parasite species, we observed highly contrasting patterns of spatial genetic structure. The frequency of close *P. falciparum* relatives (>0.50 IBD) decayed with geographic distance, and selected clonal genotypes could be linked to specific geographic subdivisions in Guyana. In *P. vivax*, significant correlation of relatedness with spatial distance did not occur among sample pairs separated by >30 km, and meaningful regional population structure could not be resolved. Limited macrographic structure in *P. vivax* may occur largely because the dormant hypnozoite liver stage unique to this species facilitates long-distance dispersal via relapse in mobile hosts. Various study results point to the influence of relapse on *P. vivax* dispersal in the study region. At health centers in Greater Georgetown, for example, an absence of travel 14 days prior to diagnosis was reported more than twice as frequently by *P. vivax* patients (178 of 1,611 (11.0%)) than by *P. falciparum* patients (45 of 867 (5.2%)). Nucleotide diversity was also significantly elevated in *P. vivax* samples representing such non-traveler cases. Given that Greater Georgetown is considered free of malaria transmission, many non-traveler *P. vivax* cases may represent reactivation of dormant parasite diversity that has accumulated in hosts over longer periods of time^39^. We also observed the recurrence of *P. vivax* clones in 11 patients for which blood spots were sequenced from separate malaria visits. These same-patient sample pairs represented symptomatic malaria episodes separated by a median of 109 days and the prescription of a 3-day Chloroquine + 14-day primaquine treatment course. They may thus not only represent direct evidence of relapse but also point to efficacy and adherence challenges relating to hypnozoicidal drugs.

Together, these observations suggest that differences in underlying parasite biology are contributing to divergent species responses to intervention, including flips from *P. falciparum* to *P. vivax* case majority observed over the last 15 years in regions nearing elimination goals (e.g., Guyana, Solomon islands, Myanmar, Cambodia, Lao PDR^1^).

In the case of *P. falciparum* in Guyana, malaria control strategy may benefit from further advancing decentralized case management and differentiating resource allocation based on real-time spatiotemporal surveillance. Similar to regions in Southeast Asia where these approaches have recently reduced *P. falciparum* cases to historic lows^40^, our study indicated a *P. falciparum* metapopulation maintained by periodic expansions from high-risk transmission areas (mines in remote riparian forest) where infrastructure is limited and occupational incentives conflict with self-care. In these settings, treatment-seeking is more likely to be delayed (increasing the risk of transmission prior to treatment) and access to regional treatment centers may involve long-distance travel through endemic terrain. It therefore may be beneficial to further decentralize medical capacity from regional treatment centers to localities with strong outbreak signals (e.g., Puruni Landing) or to strategic points along travel routes that may facilitate parasite dispersal (e.g., along the Mazaruni River, a possible conduit for clonal *P. falciparum* dispersal based on this study). Self-administered diagnostic and treatment approaches (e.g., Malakit^41^) might additionally help cover remote areas, with monitoring to evaluate the extent to which incorrect self-treatment may negatively impact user health or contribute to resistance emergence.

In the case of *P. vivax* control, our study suggests that similar strategies of decentralized and focally intensified intervention will be less efficacious. We do not observe tractable metapopulation structure that would help prioritize intervention foci, and various results point to the risk of latent dispersal processes undermining geographically differentiated intervention. Decentralizing treatment resources, especially the integration of unsupervised treatment kits, also appear less tractable due to the greater complexity of safe and effective ‘radical’ *P. vivax* cure (treatment targeting both bloodstage and hypnozoite parasites). Hypnozoite clearance generally involves two-week treatment with 8-aminoquinolone drugs and requires prior glucose-6-phosphate dehydrogenase (G6PD) deficiency assessment of haemolysis risk^15^. This also makes conventional mass drug administration approaches targeting latent *P. vivax* reservoirs difficult to realize in settings without exceptionally low rates of G6PD deficiency^42^ and exceptionally consistent treatment compliance^43^. It is therefore necessary to advance work on alternative implementation strategies (e.g., CUREMA^44^ approaches to radical cure in medically underserved populations of the Guiana Shield) and towards the discovery of new hypnozoicidal drugs.

In the interim, fortifying preventive vector control and risk awareness programs is also important. A recent study suggests that increased risk perception is associated with increased preventive behavior against vector borne disease in Guyana, although effect size is small (4-5%)^45^. Effect size might be increased if educational programs more strongly convey risk of onward malaria transmission as opposed to risk solely towards personal health. Such altruistic risk perception may be relevant to the more rapid, pre-symptomatic spread of *P. vivax* malaria into local vectors, including upon miner return to non-mining home environments.

This study had several limitations. Importantly, the near-relative detection approach we used to describe parasite population structure focuses on outlier IBD relationships and therefore underuses available sequence content. We also did not infer IBD relationships involving polyclonal infections, which constituted 17.9% of our genomic sample set. Furthermore, genomic sample collection relied on a sentinel method which may bias against parasite diversity found in remote, disconnected transmission cycles (e.g., Amerindian settlements). It was also challenging to address the transitory nature of human settlement in the hinterlands of Guyana, where conventional population sizes are not recorded or do not apply. This primarily complicated the estimation of zonal incidence and its relationship to polyclonality rates. High host mobility in undercharted hinterland areas also added uncertainty to geographic infection source estimates.

Despite these limitations, this study clearly exposes the discrepant profiles of co-endemic *P. falciparum* and *P. vivax* malaria in a low transmission setting, and emphasizes the need for distinct intervention and surveillance approaches in order to succeed in eliminating both species. The current disproportionate investment in drugs, vaccines, and monoclonal antibodies for *P. falciparum* increases the likelihood that *P. vivax* will persist in many co-endemic regions after *P. falciparum* is eliminated, forestalling the ultimate goal of eliminating malaria from all regions and demographics.

## Methods

### Epidemiological database and definitions/categorizations

The epidemiological database analyzed in this study comprised all malaria episodes detected in Guyana via passive surveillance (estimated to represent approximately 80% of cases^1^) and reported to the National Malaria Program between 2007 and 2019. Malaria episodes were defined using both medical diagnoses and parasitological tests (blood microscopy, and less commonly, rapid diagnostic tests). Patient records were curated by the Vector Control Services (VCS) division of the GMOH and anonymized records of patient age, gender, nationality, and self-reported ethnicity (categorized as “Afroguyanese”, “Amerindian”, “East Indian”, “Chinese”, “Portuguese”, “European”, “Mixed”, or “Other”) were provided to the Harvard T.H. Chan School of Public Health (HSPH) following approval by the Harvard University Area Human Research Protection Program (protocol IRB18-1638) and by ethical committees of the GMOH. Infection localities were inferred based on voluntary responses about *“Where [the] patient stayed 2 weeks ago”* (see survey form in Supplementary Fig. 17). Each travel history response was classified to one of 460 country-wide localities with known latitude/longitude coordinates.

To define epidemiological zones, we mapped malaria survey sites used by the GMOH (https://gazetteer.glsc.gov.gy/gazetteer/#7/3.650/-57.129) onto a custom shape file containing Guyana#s primary river and road coordinates and onto a motorized transport resistance raster obtained from https://malariaatlas.org/ (Supplementary Fig. 2). Sites were clustered based on river/road connectivity in the R package RIVERDIST^46^ v0.16.3, travel conductance using the R package GDISTANCE^47^ v1.6.4, and manual assessment of river/road and resistance layers in QGIS^48^ v2.18.4. Transmission intensity was estimated for each epidemiological zone using population size estimates from the LandScan Project^49^. The LandScan approach uses dasymetric modeling to disaggregate census counts based on remotely sensed images^49^. We used Landscan Global 2019 data to estimate average population density within 10 arcmin (ca. 20 km) of the centroid representing all localities contributing malaria cases within an epidemiological zone. We then divided the zonal case count by its LandScan population density estimate.

### Clinical sample material, sequencing, and variant calling

Following informed consent to analyze parasite genetic polymorphism using patient blood, samples were collected from microscopy or RDT-positive patients by spotting approximately 50-200 μl whole blood onto Whatman FTA filter paper cards. Collection was carried out by VCS between January 2020 and June 2021 at medical facilities in Bartica, Georgetown, Mahdia, Lethem, and Port Kaituma, Guyana. Samples were stored at room temperature using individual desiccant packets before shipment to HSPH. The same anonymized patient metadata variables as described in the previous section (Supplementary Fig. 17) were also provided to HSPH. An additional 23 dried blood spot samples were collected in Venezuela at the Institute of Tropical Medicine, Central University of Venezuela (Caracas, bioethics permit CEC-IMT 12/2013) and Biomedical Research and Therapeutic Vaccines Institute (Ciudad Bolívar, bioethics permit CHURPCBBS-008-2019) in 2019. Metadata for these samples (collection date and reported place of stay (municipality) two weeks prior to diagnosis) was likewise de-identified prior to HSPH access (Supplementary Table 2).

We extracted total genomic DNA from dried blood spot samples (approximately 20-35 mm^2^ spotted area punched per sample) using KingFisher Ready DNA Ultra 2.0 Prefilled Plates on the Kingfisher Flex instrument (ThermoFisher Scientific). We subsequently applied whole-genome amplification to DNA extracts using sets of ten *P. falciparum*^50^ and/or *P. vivax*-selective oligos^44^ (set selection based on previous microscopy or RDT-based species assignment) and used AMPure XP magnetic beads (Beckman Coulter) to exchange post-reaction sample buffer to 10 mM Tris-HCl + 0.1 mM EDTA. Final library construction using the NEBNext Ultra II FS DNA Library Prep Kit (NEB #E6177) and 2 x 150 bp sequencing on the Illumina HiSeqX platform (150-bp paired-end reads) was completed at the Broad Institute. We aligned reads to the *P. falciparum* 3D7 and *P. vivax* PvP01 reference genome assemblies using BWA-MEM^52^ and called SNPs and INDELs using GATK^53^ v3.5-0-g36282e4 ‘HaplotypeCaller’ and ‘GenotypeGVCFs’ according to best practices defined by the Pf3k consortium (http://www.malariagen.net/data_package/pf3k-5/). The mapping and joint variant call process also included sample accessions ERR1818176 and ERR2496572 representing malaria infections from Venezuela (further geographic specifics unknown). All downstream genetic analysis focused on SNP sites in core regions of the genome^54,55^ and >5 bp from any INDEL call. The base SNP call set created for each species also excluded sites at which >7.5% of monoclonal samples showed multi-allelic (‘heterozygous’) calls.

### Parasite genetic analyses

We assessed polyclonality using THEREALMcCOIL^34^ v2, a Bayesian Markov chain Monte Carlo approach that simultaneously estimates population allele frequency for each SNP and COI for each individual in the sample set. We applied the categorical method (classifying SNP calls binarily as homozygous or heterozygous regardless of the signal intensity of each component allele) to bi-allelic SNP sites with ≥10% minor allele frequency and ≥90% call success across samples. We further subsampled input to limit computational costs. Subsampling using the ‘-- thin’ function in VCFtools^56^ v0.1.15 resulted in 337 *P. falciparum* and 350 *P. vivax* input sites. We kept input parameters in default and used the lower bound (quantile = 2.5%) output value as a conservative estimate of sample COI.

Analyses of genetic diversity and relatedness only included monoclonal samples. We estimated nucleotide diversity from the base SNP call sets (previous section) by measuring average nucleotide differences over discrete 100 kbp windows (no overlap) with VCFtools^56^ v0.1.15. We measured linkage decay over bi-allelic SNP sites by recoding calls to non-reference allele counts (0, 1, or 2) and computing linkage (r^2^) in sliding 10 kbp windows (200 bp steps) in PLINK^57^ v1.90b1g.

We estimated genetic relatedness between samples based on the concept of IBD. Unlike identity by state (IBS), IBD specifically represents genetic similarity inferred to have been inherited from a common ancestor based on factors affecting linkage likelihood such as population allele frequency, recombination rate, and chromosomal distances between variant sites. IBD generally enhances representation of recent ancestry in obligately sexual species in which divergence history is influenced more strongly by recombination than by the individual accumulation of point mutation events. We ran hmmIBD^33^ in default settings to estimate population allele frequency and infer presence or absence of IBD at all bi-allelic SNPs with ≥2% minor allele frequency and ≥70% call success across samples. We used the fraction of all variant sites inferred as IBD (‘fract_sites_IBD’) to summarize relatedness for each sample pair.

We used the R package IGRAPH^58^ v1.3.5 to generate connected graphs (clusters) in which edges represent sample pairs with relatedness values above a specified threshold. Clusters therefore represent groups of samples (nodes) in which each sample is connected by at least one direct edge to another sample (but simultaneous direct connections to additional cluster members are not required).

We also correlated relatedness values to Euclidean distances calculated from latitude and longitude (WGS 84) coordinates projected onto a common xy plane (EPSG 3786). We used Mantel tests to assess statistical significance between genetic and spatial distance matrices using the R package VEGAN^59^ v2.6-4. We applied repeated random draws of 10 samples from each epidemiological zone (drawing until depletion without replacement) to build each distance matrix. We also mapped the frequency of pairwise relatedness between epidemiological zones using shape files from the Natural Earth public domain map dataset^60^.

Finally, we also assessed the possibility that cases of clonal IBD detection in *P. vivax* represent repeated blood spot sampling events from the same patient (e.g., a first clinical visit due to primary infection and one or more later clinical visits due to relapse or recrudescence by the same parasite genotype). HSPH authors sent a query list of 705 *P. vivax* samples to the GMOH and the GMOH flagged each according to whether it represents a patient which is also represented by another sample in the query list. The list returned by the GMOH indicated 1x (i.e., non-repeat) patient representation for 589 samples, 2x patient representation for 92 samples (necessarily representing 46 patients) and ≥3x patient representation for 22 samples (necessarily representing ≤7 patients). The returned list did not specify which sample sets corresponded to the same patient.

## Data Availability

All data produced in the present study are available upon reasonable request to the authors.

## Acknowledgements

We thank the participants who contributed blood samples to the study, as well as the technicians who collected and processed the samples. This study was supported by the Bill & Melinda Gates Foundation (INV-009416). Under the grant conditions of the Foundation, a Creative Commons Attribution 4.0 Generic License has been assigned to the Author Accepted Manuscript version that might arise from this submission. This study was also supported with federal funds from the National Institute of Allergy and Infectious Diseases, National Institutes of Health, Department of Health and Human Services, under Grant Number U19AI110818 to the Broad Institute. Sampling in Venezuela was additionally supported by the Scottish Funding Council Global Challenges Research Fund (GCRF) Small Grants Fund SFC/AN/12/2017 and the GCRF Vector-borne Disease Control Network (EP/T003782/1).

## Data Availability Statement

The sequence data generated by this study have been deposited in the NCBI Sequence Read Archive under BioProject PRJNA809659.

## Supplementary Text 1

### Relationships between samples collected in Guyana and Venezuela

We assessed whether our small comparator set from Venezuela (22 monoclonal samples) could indicate the presence of cross-border parasite genetic divergence and transmission of imported strains within Guyana. The Venezuelan comparator set primarily represents infections from the eastern states of Bolívar and Sucre. Two *P. falciparum* samples (PW0065-C and SPT26229) and one *P. vivax* sample (CEM541_Pv-9) lack travel history data. In *P. falciparum*, the IBD distribution for sample-pairs representing comparisons within Guyana (median = 0.283) appeared right-shifted (Wilcoxon test, W= 57691375, p < 0.001) relative to the IBD distribution for sample-pairs representing cross-border comparisons (median = 0.261, Supplementary Fig. 9a). To further visualize this divergence signal in *P. falciparum* and to screen for imported transmission, we mapped the highest IBD value observed for each sample with respect to samples representing Venezuela (‘MaxVZ’, Supplementary Fig. 9b). Only three infections from Guyana showed aberrant (>2 sd above average) maxVZ. Two of these three (G4G410 and G4G1043) were clonal (>0.90 IBD) with respect to Venezuelan infections Venez_001_F1 and PW0065-C (Supplementary Fig. 9c,d). Highly aberrant (> 3 sd above average) IBD relative to G4G410 or G4G1043 occurred in five of nine Venezuelan infections but in just one of 527 Guyanese samples. These observations suggest that G4G410 and G4G1043 represent cases of imported transmission from Venezuela into Guyana. Observing just two such cases in the sample set suggests that imported transmission is infrequent (despite health posts near the border frequently experiencing foreign cases – see cyan pie slices in Supplementary Fig. 12) but verification is required with larger Venezuelan sample sets.

In *P. vivax,* G-G and G-V distributions were also statistically distinguishable (p<0.001) but were very close in absolute overlap (median 0.046 vs. 0.041 (respectively) Supplementary Fig. 9e)). Although scarce, aberrant maxVZ values (>2 sd above average) occurred within Guyana more frequently in *P. vivax* (7/474) than in *P. falciparum* (3/528) and occurred within Venezuela less frequently in *P. vivax* (5/13) than in *P. falciparum* (6/9) (Supplementary Fig. 9f vs. Supplementary Fig. 9b).

Principal component analysis using direct SNP data (IBS instead of IBD, Supplementary Fig. 9g) did not help clarify Guyana-Venezuela population patterns in either species. Maps of alternative IBD features (e.g., maximum intrachromosomal IBD tract length) were also difficult to interpret (Supplementary Fig. 9h-m).

---

**Supplementary Fig. 1.**
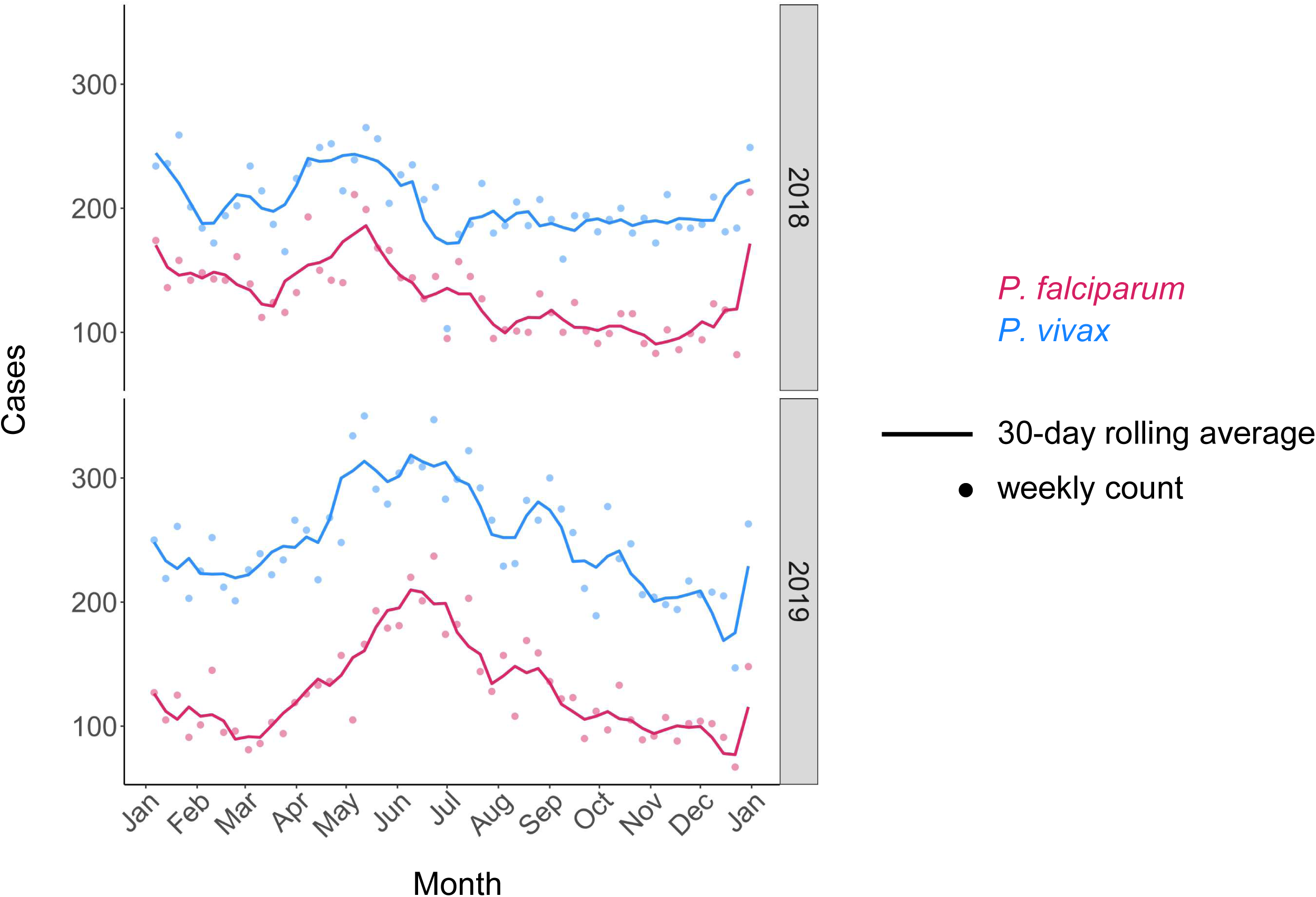
Intra-annual variation in *P. falciparum* and *P. vivax* cases in Guyana. Points represent weekly reported cases and lines represent 30-day rolling averages in 2018 and 2019.

**Supplementary Fig. 2.**
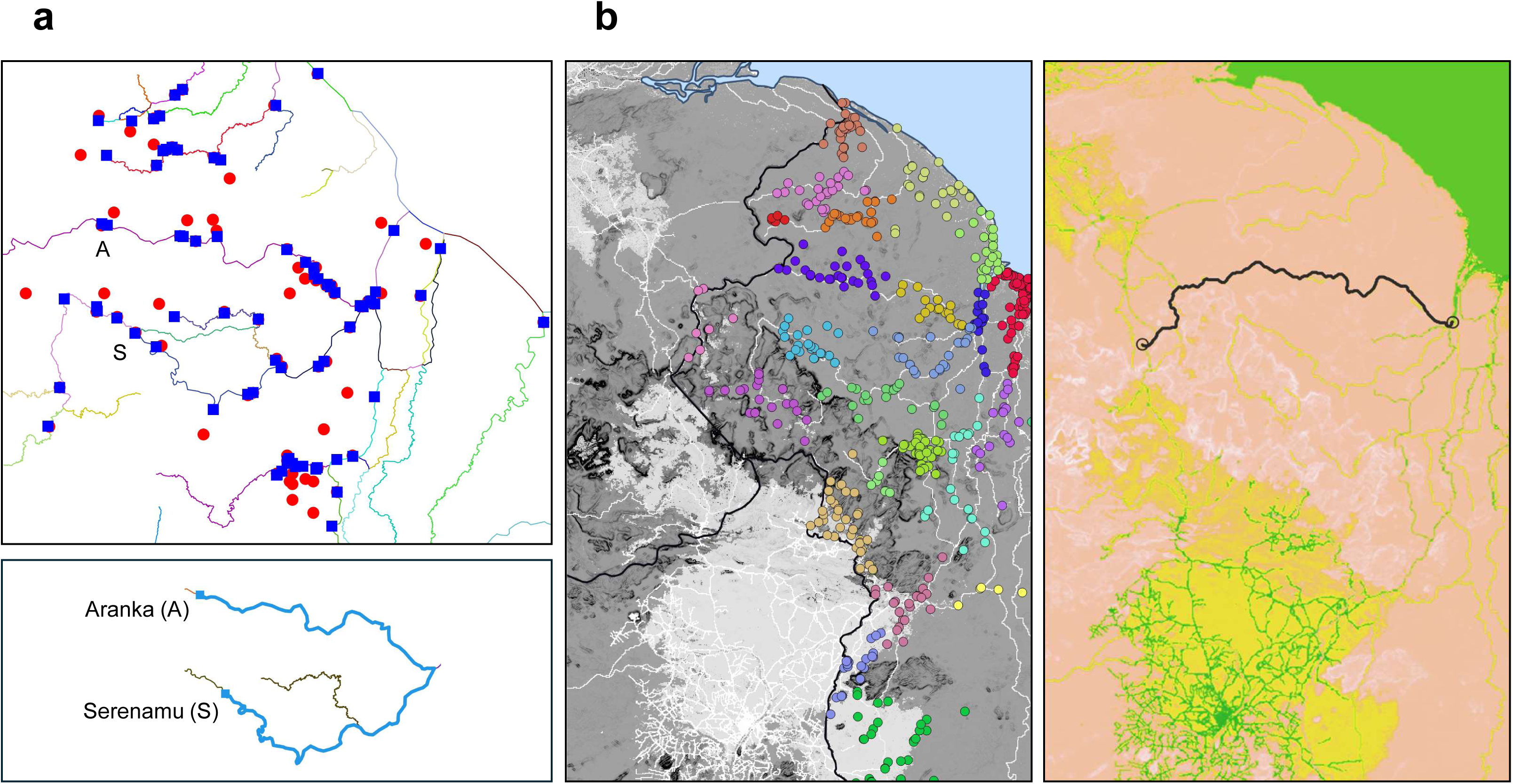
Epidemiological zoning based on connectivity analysis in Guyana. Diagnosis and infection localities (inferred based on reported stay 2 weeks prior to diagnosis) were grouped into 27 epidemiological zones. Though not automated using any fixed thresholds, grouping decisions were partially informed by river distance calculation and resistance surface analysis. a) To measure river distances, localities were first fitted to nearest river coordinates (red and blue points, respectively). Distances along rivers were then measured between assigned coordinates (e.g., Aranka to Serenamu, bottom plot). b) For resistance surface analysis, localities were plotted on a mobility friction map obtained from https://malariaatlas.org/. Light grey and white represent highest conductance to motorized travel. The right plot illustrates how a least cost path (black line) inferred for Las Cristinas (Venezuela) to Bartica (Guyana)) matches the flow of the Cuyuni River. Yellow and green represent highest conductance to motorized travel.

**Supplementary Fig. 3.**
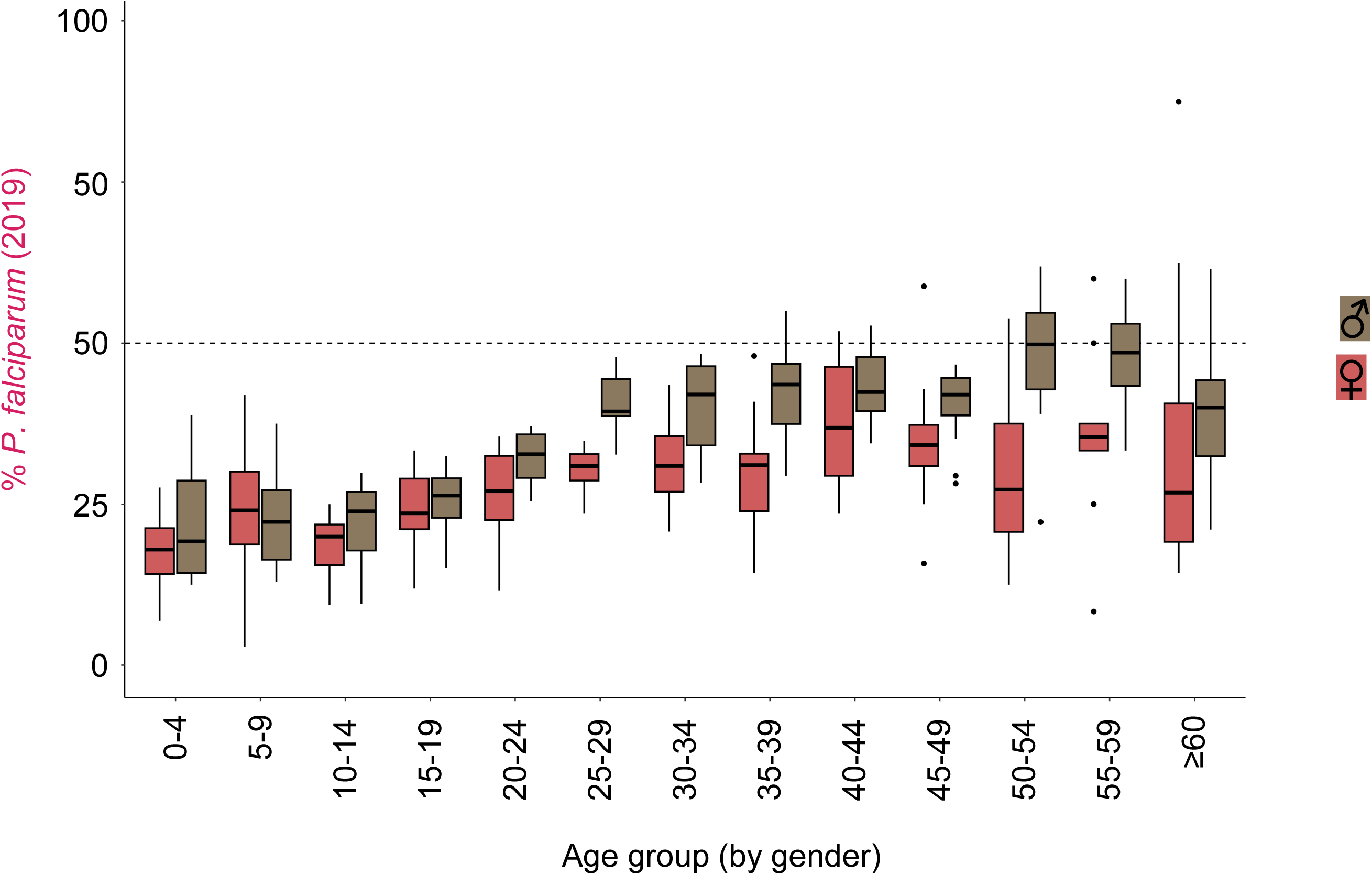
Relative *P. falciparum* prevalence with respect to age and gender in Guyana. Boxplots summarize monthly variation (median and quartiles) in the percent of malaria cases representing *P. falciparum* in 2019. Brown and red represent values for male and female cases, respectively.

**Supplementary Fig. 4.**
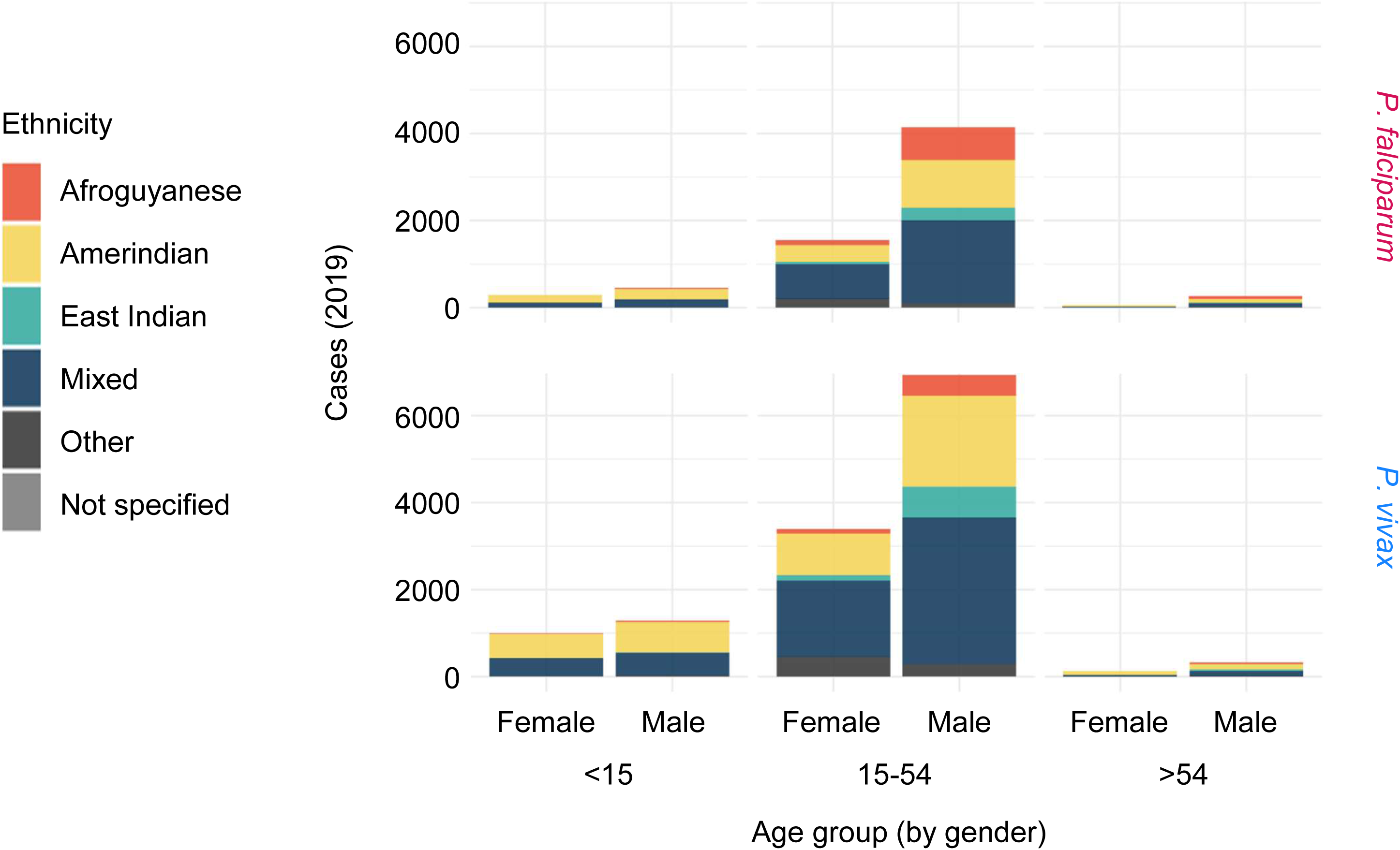
*P. falciparum* and *P. vivax* case counts be age, gender, and ethnicity in Guyana. Colors in stacked bar charts indicate self-reported ethnicities (key at left) among malaria patients in 2019.

**Supplementary Fig. 5.**
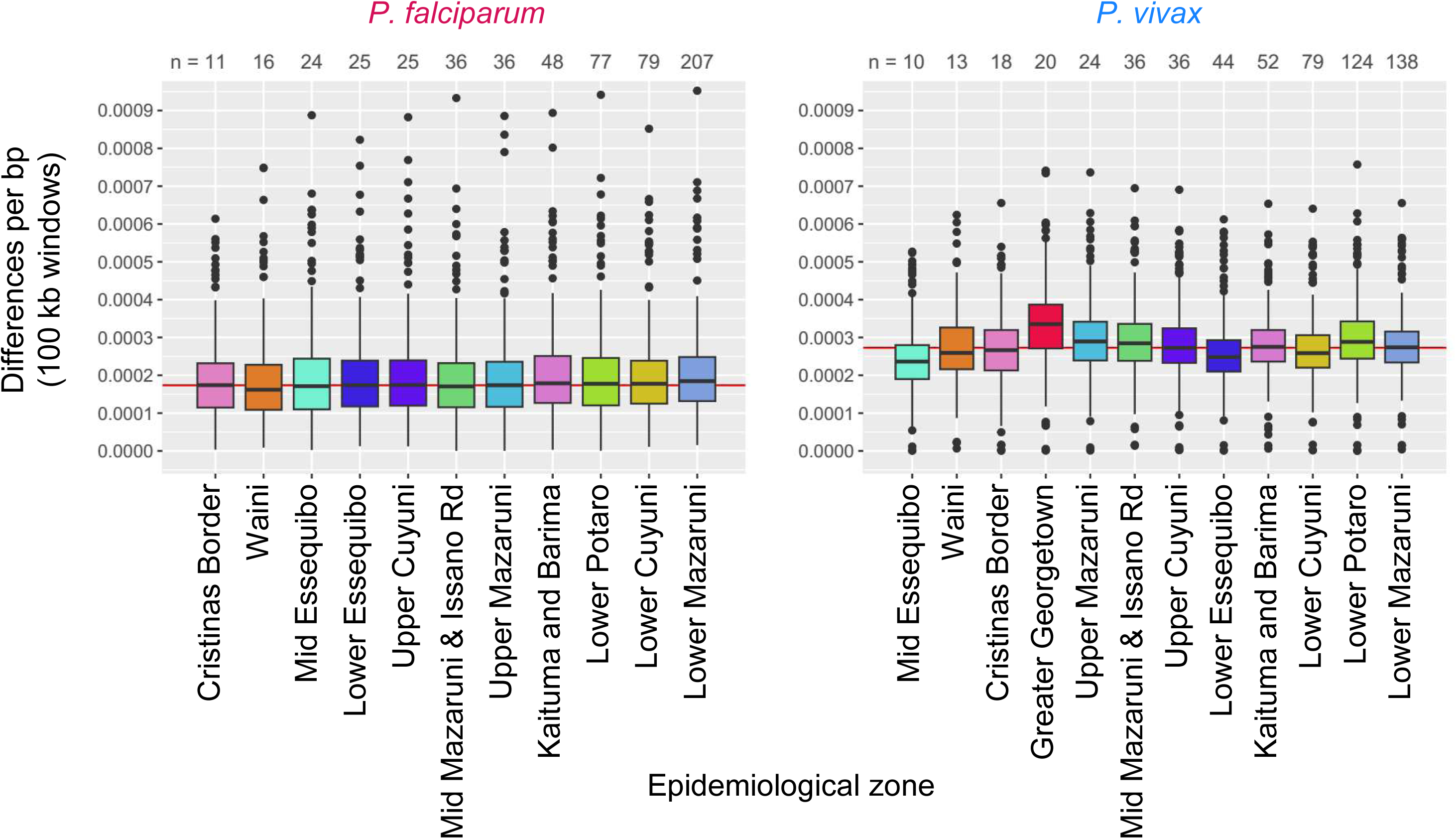
*P. falciparum* and *P. vivax* pairwise nucleotide diversity among epidemiological zones in Guyana. Boxplots summarize variation (median and quartiles) in windowed π values (y-axis) for each epidemiological zone (x-axis) represented by ≥ 10 genomic samples (see top) in 2020-21.

**Supplementary Fig. 6.**
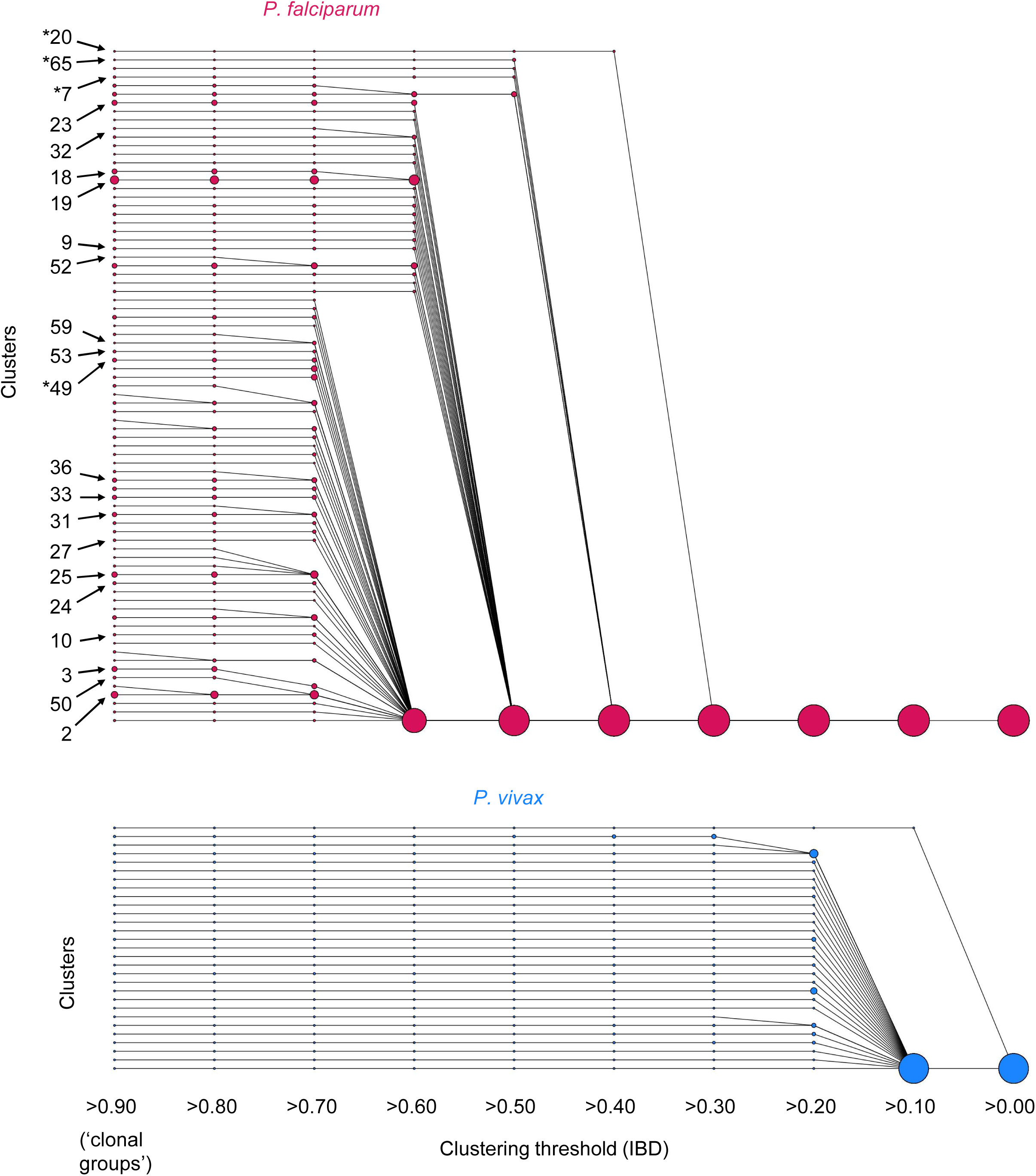
*P. falciparum* and *P. vivax* clustering relationships in Guyana and Venezuela. Clusters (circles) represent groups of samples in which each sample is related to at least one other sample in the group at the specified IBD threshold (x-axis, decreasing from left). Circle sizes represent cluster membership sizes, minimally two. Clusters adjacent on the x-axis are connected by a line if the right (lower threshold) cluster contains all the members of the left (higher threshold) cluster. Arrows at left indicate *P. falciparum* clonal group IDs mapped in Supplementary Fig. 8. Analysis includes samples from Guyana (2020-21) and Venezuela (2015-16 and 2019).

**Supplementary Fig. 7.**
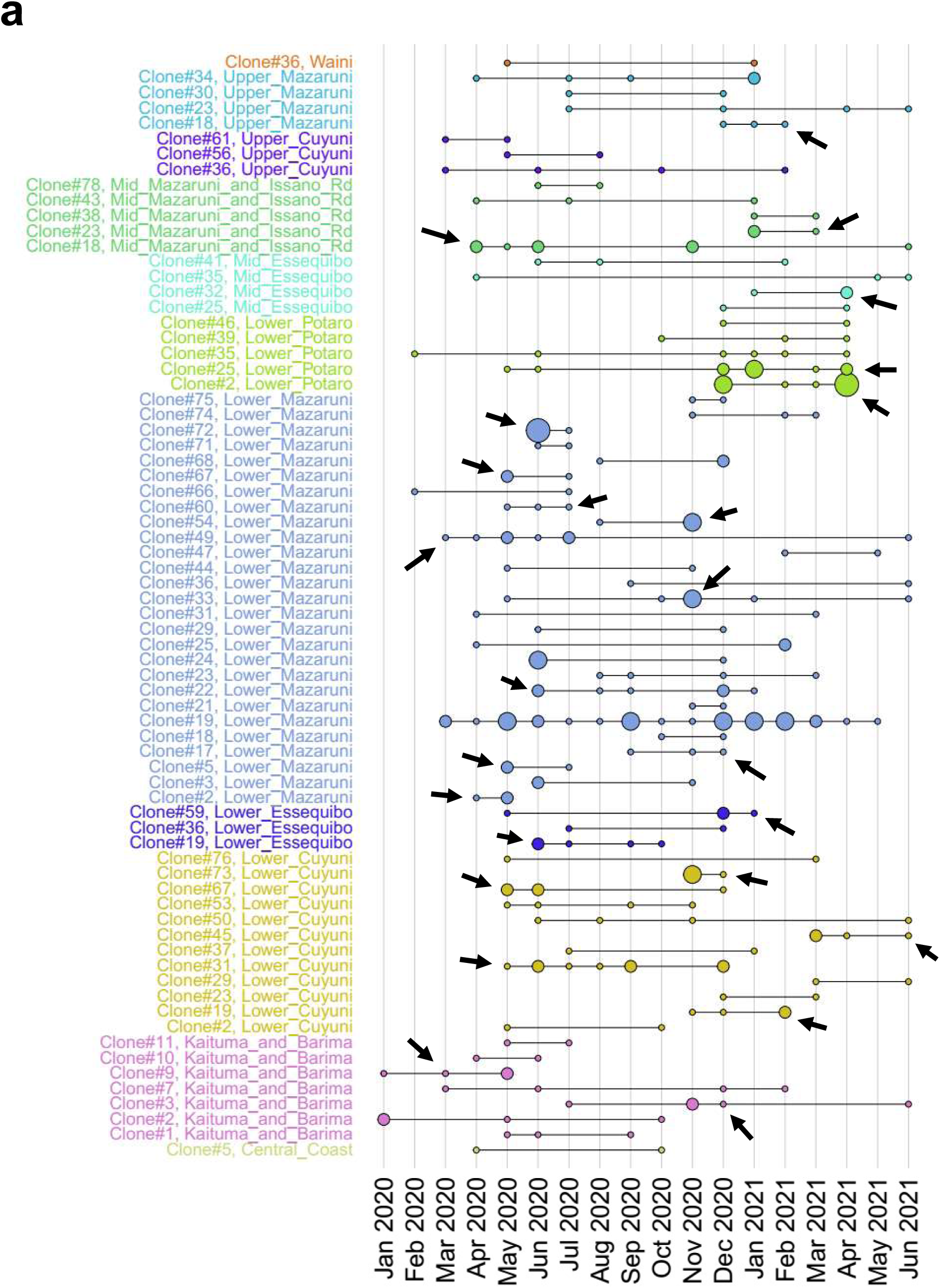
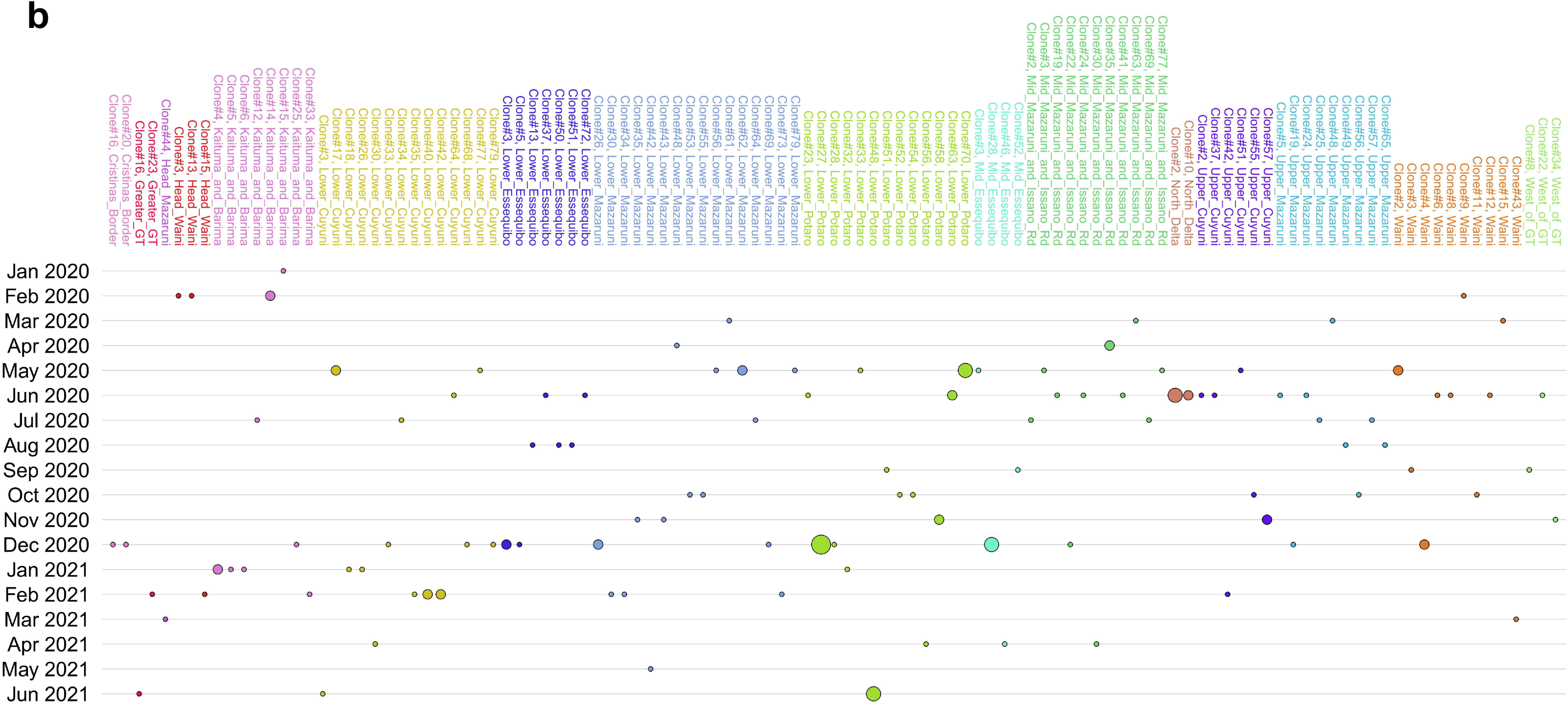
Spatiotemporal variation in *P. falciparum* clonal group membership in Guyana. The y-axis lists the clonal groups detected in each epidemiological zone (colors) in 2020-21. Group detection (circle size represents the number of group members detected) is plotted by month on the x-axis. **a)** When detections occur in multiple months per zone, these are connected by black lines. Arrows indicate instances where detection is temporally aggregated, i.e., a 4 month window contains ≥3 group members and also represents >50% of all members within that zone. **b)** In the majority of zones, membership detection does not occur in multiple months. Plots are split into two pages using flipped axes only to fit figure dimensions.

**Supplementary Fig. 8.**
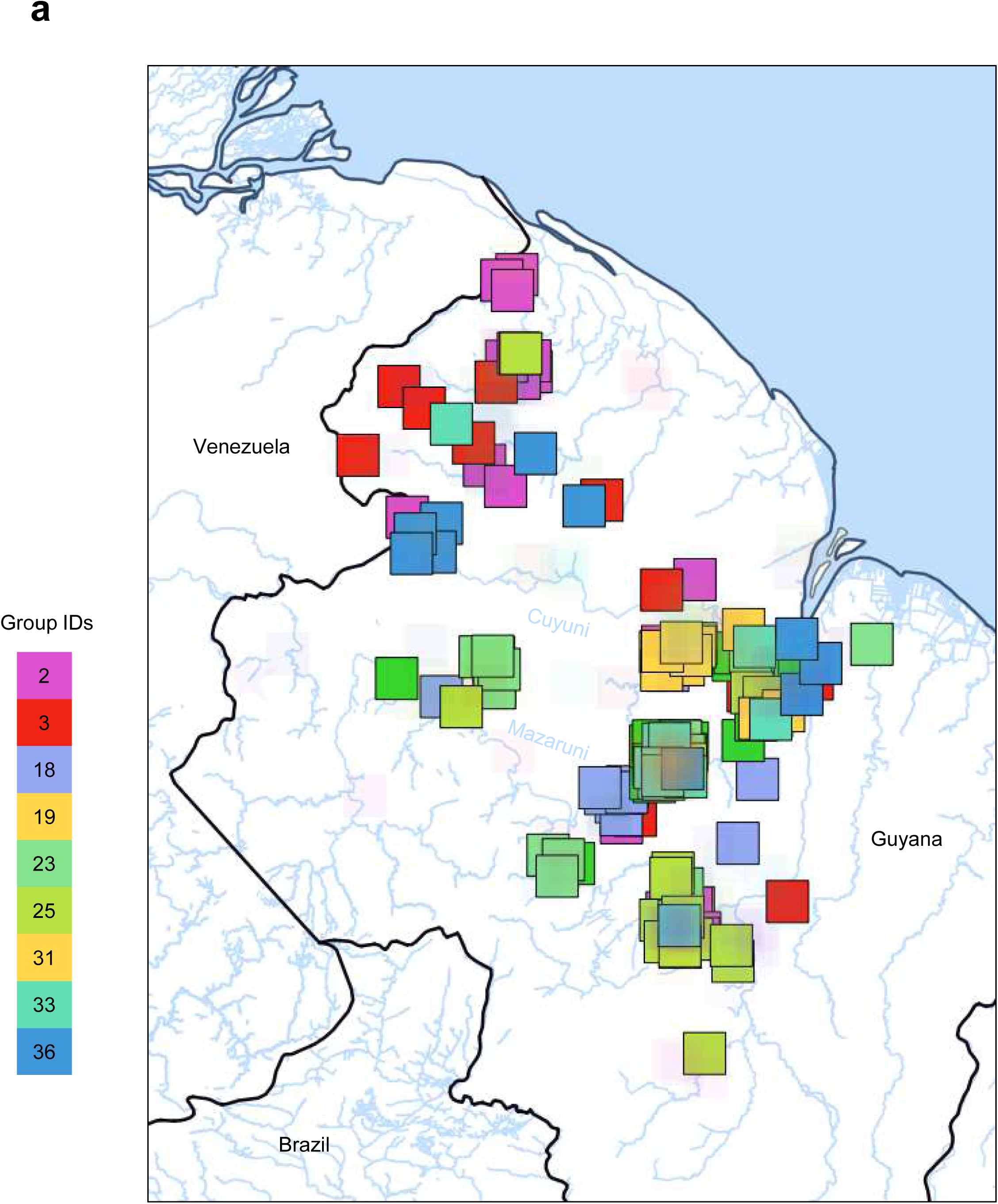
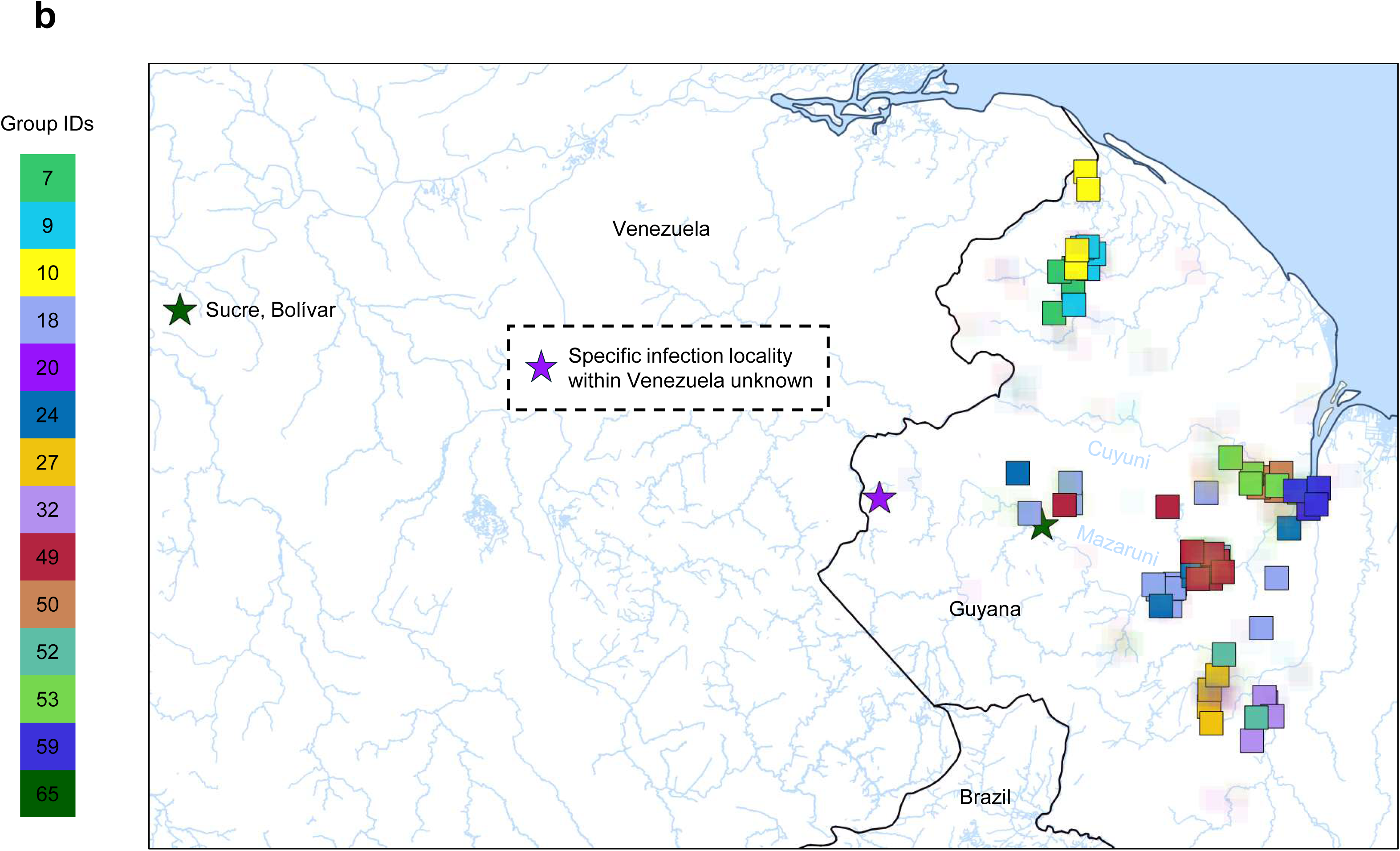
*P. falciparum* clonal group mapping in Guyana and Venezuela. a) Large clonal groups (≥10 members each) in 2020-21. **b)** Selected clonal groups highlighted in main text (2015, 2019, and 2020-21). Colors indicate group IDs (key at top left). Star symbols indicate clonal groups with partial membership in Venezuela (#65 in Sucre municipality (2019), Bolívar state, and #20, specific locality unknown (2015)). Semi-transparent symbols indicate the presence of other clonal groups aside than those being highlighted.

**Supplementary Fig. 9.**
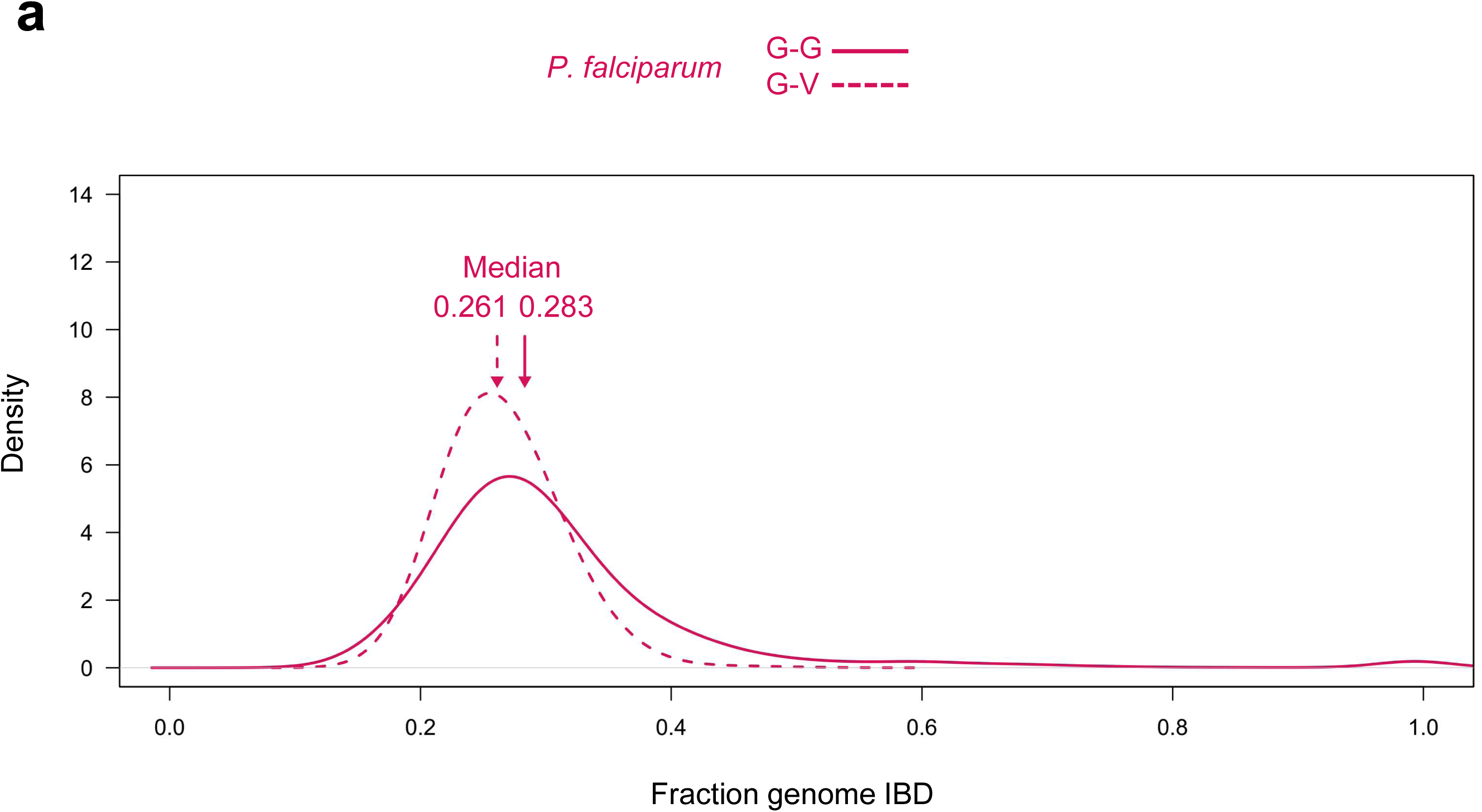
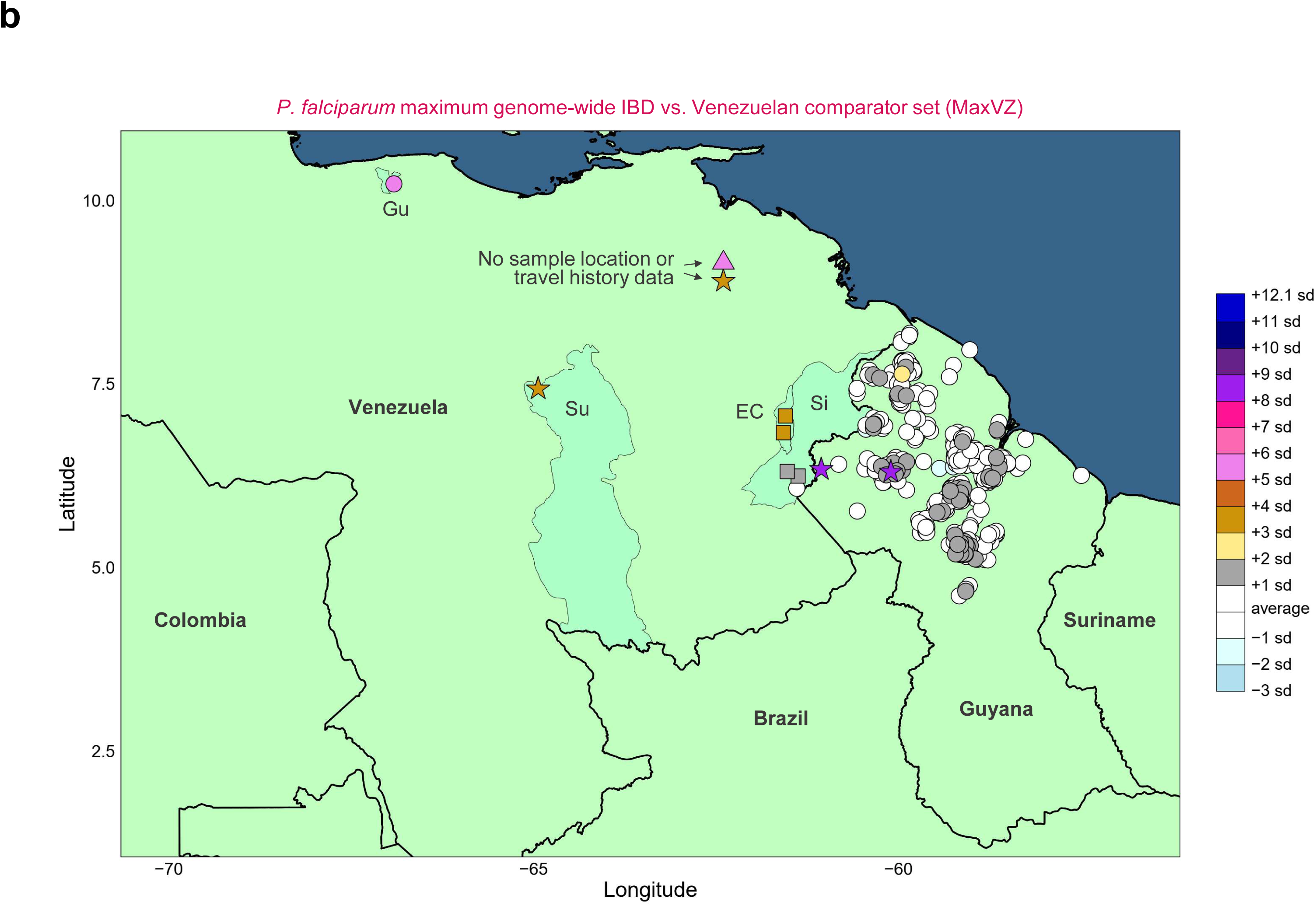
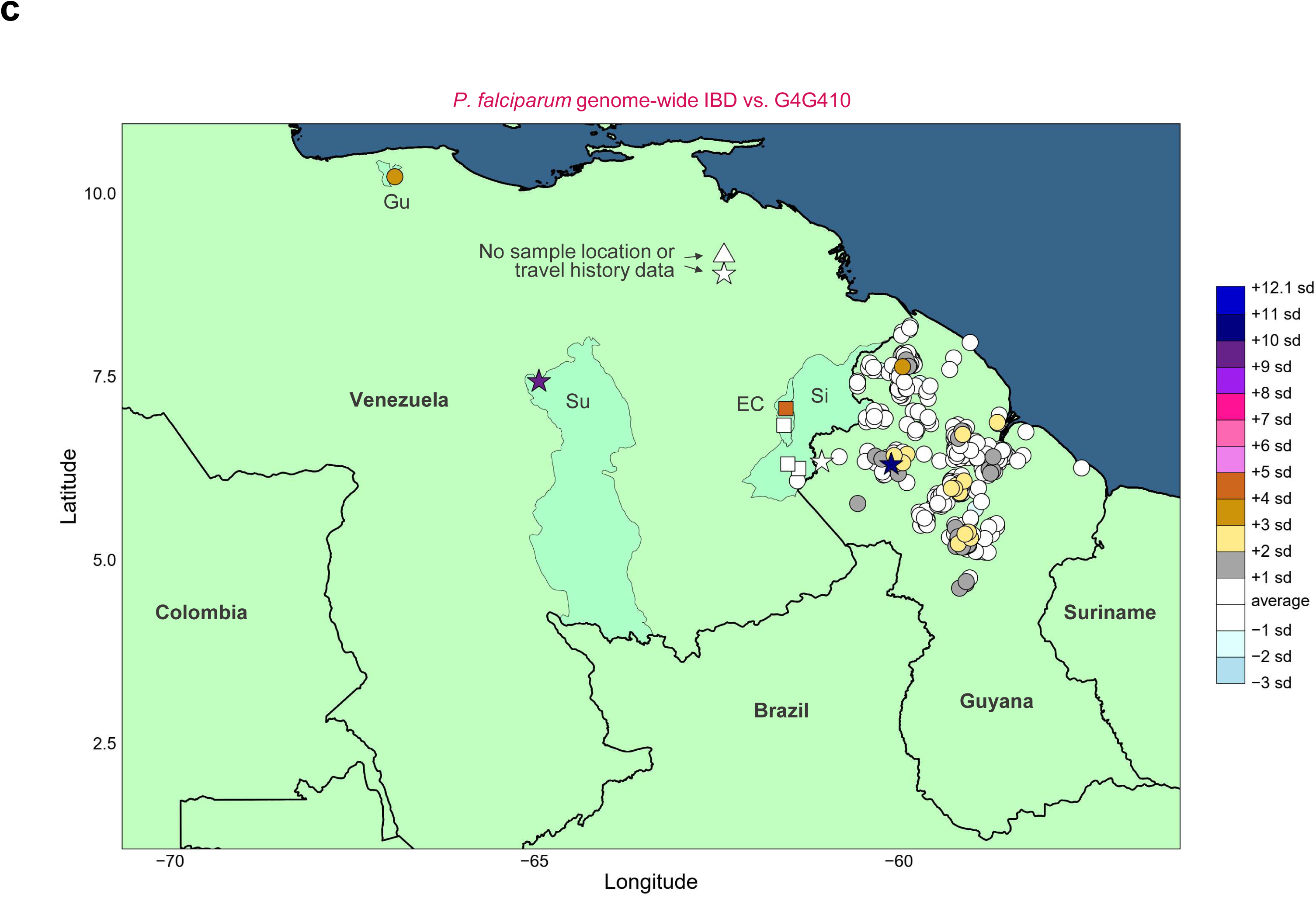
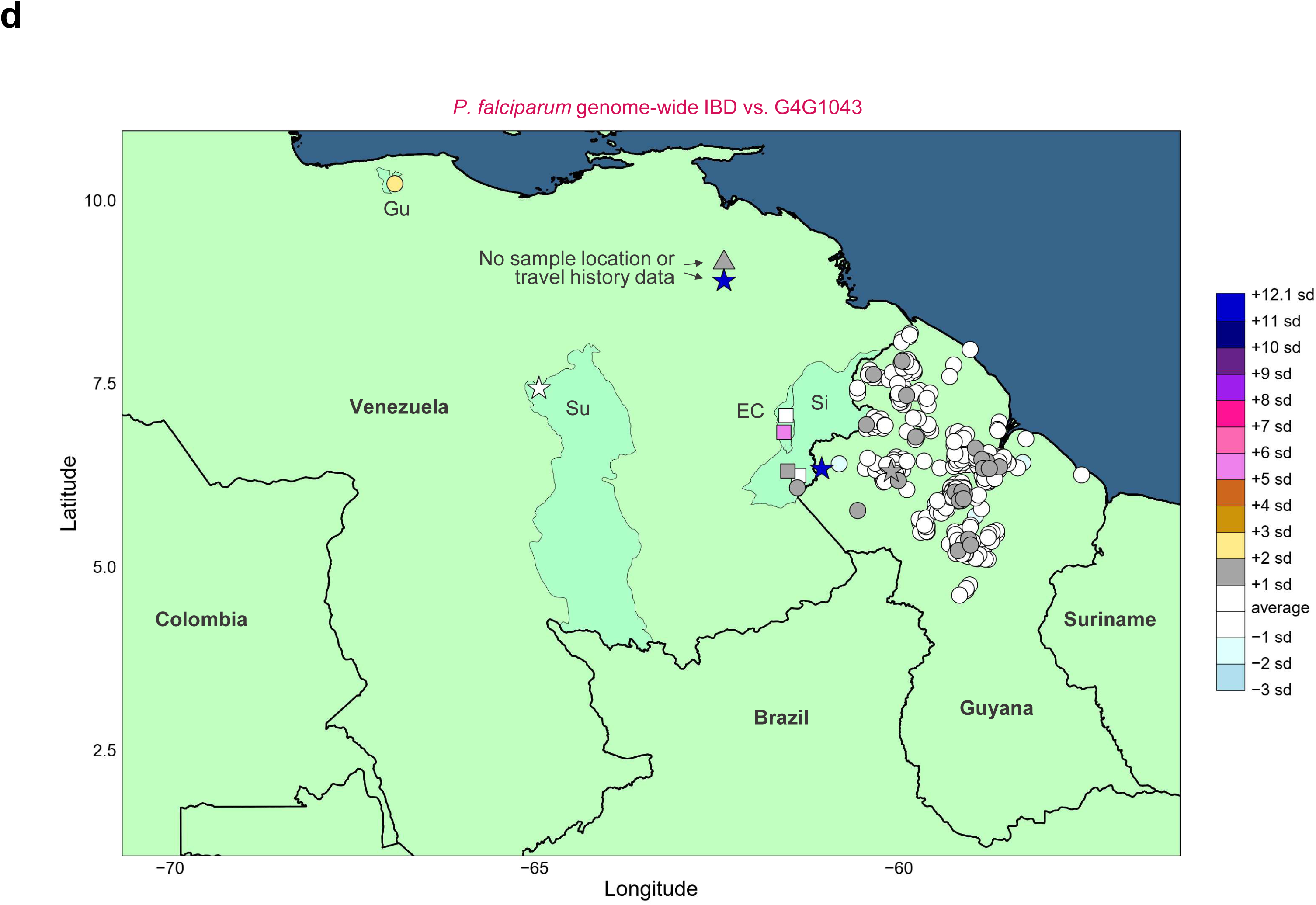
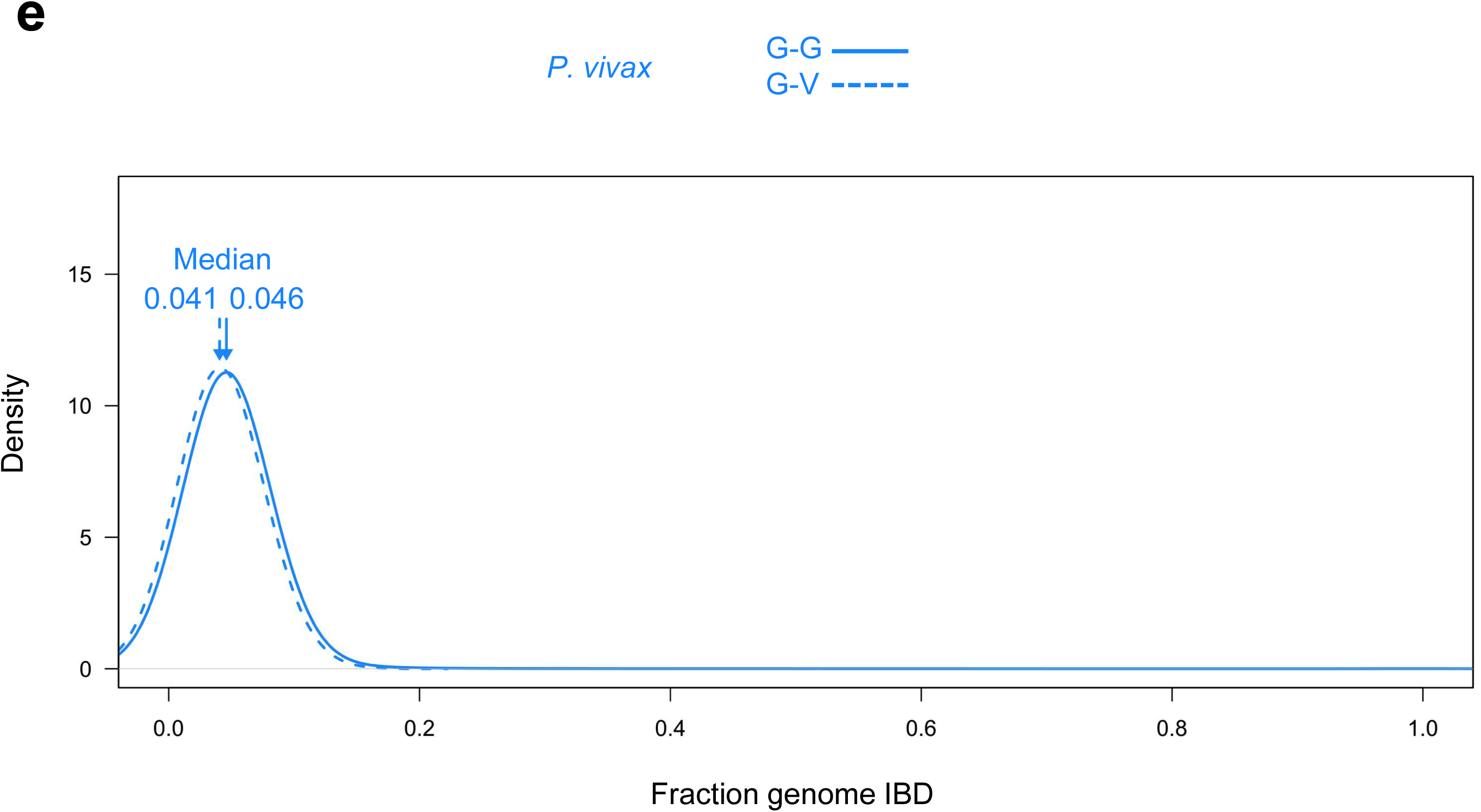
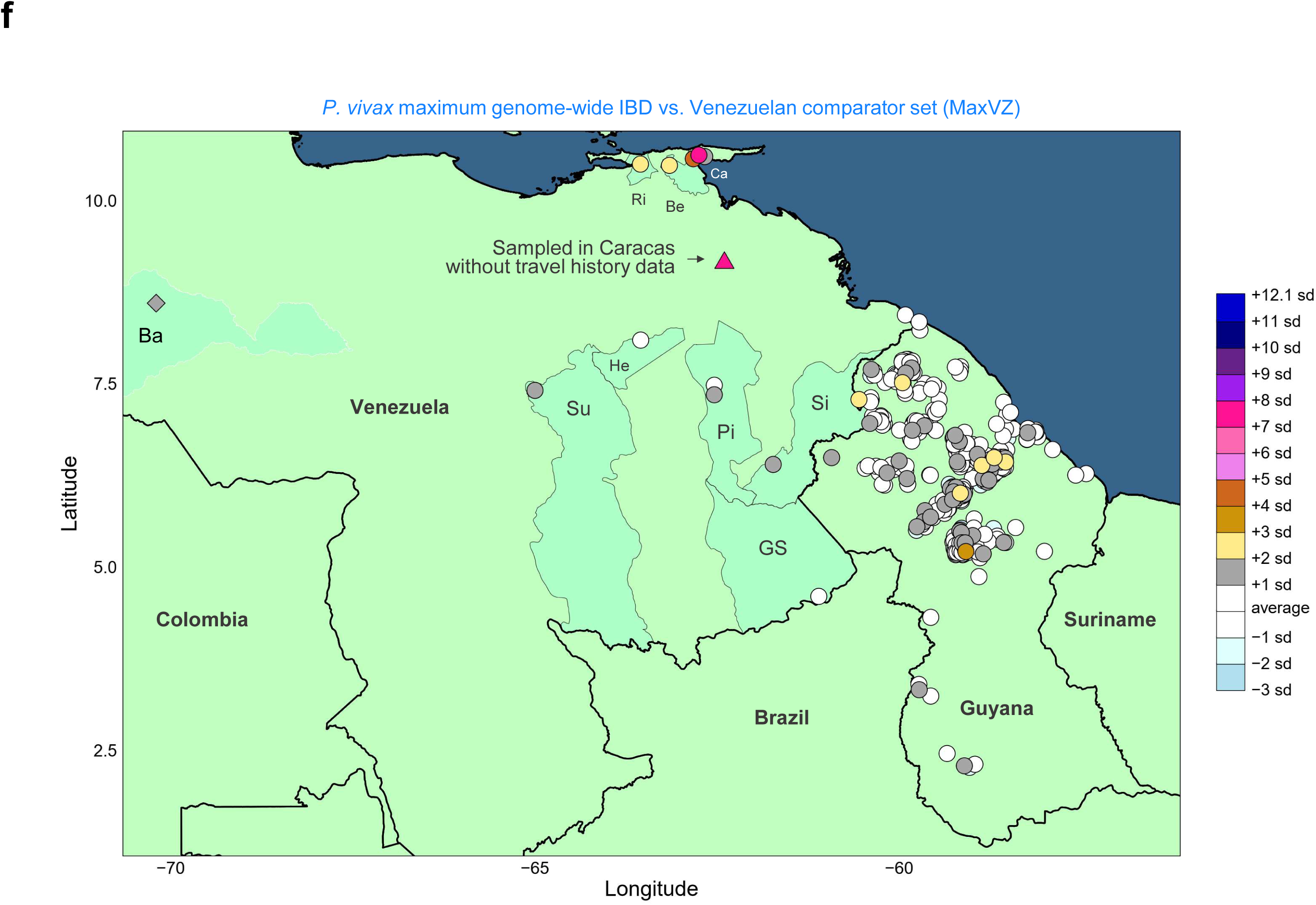
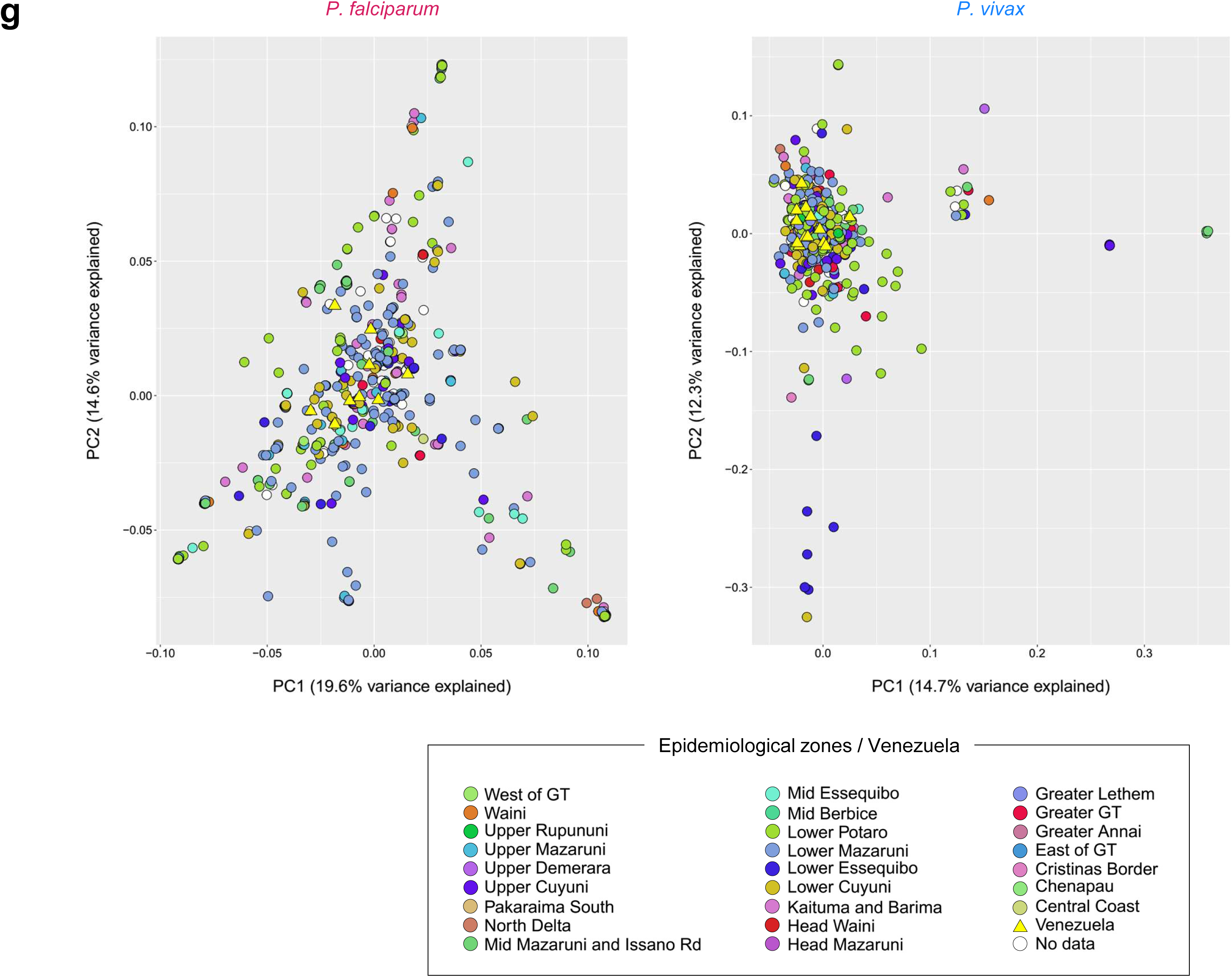
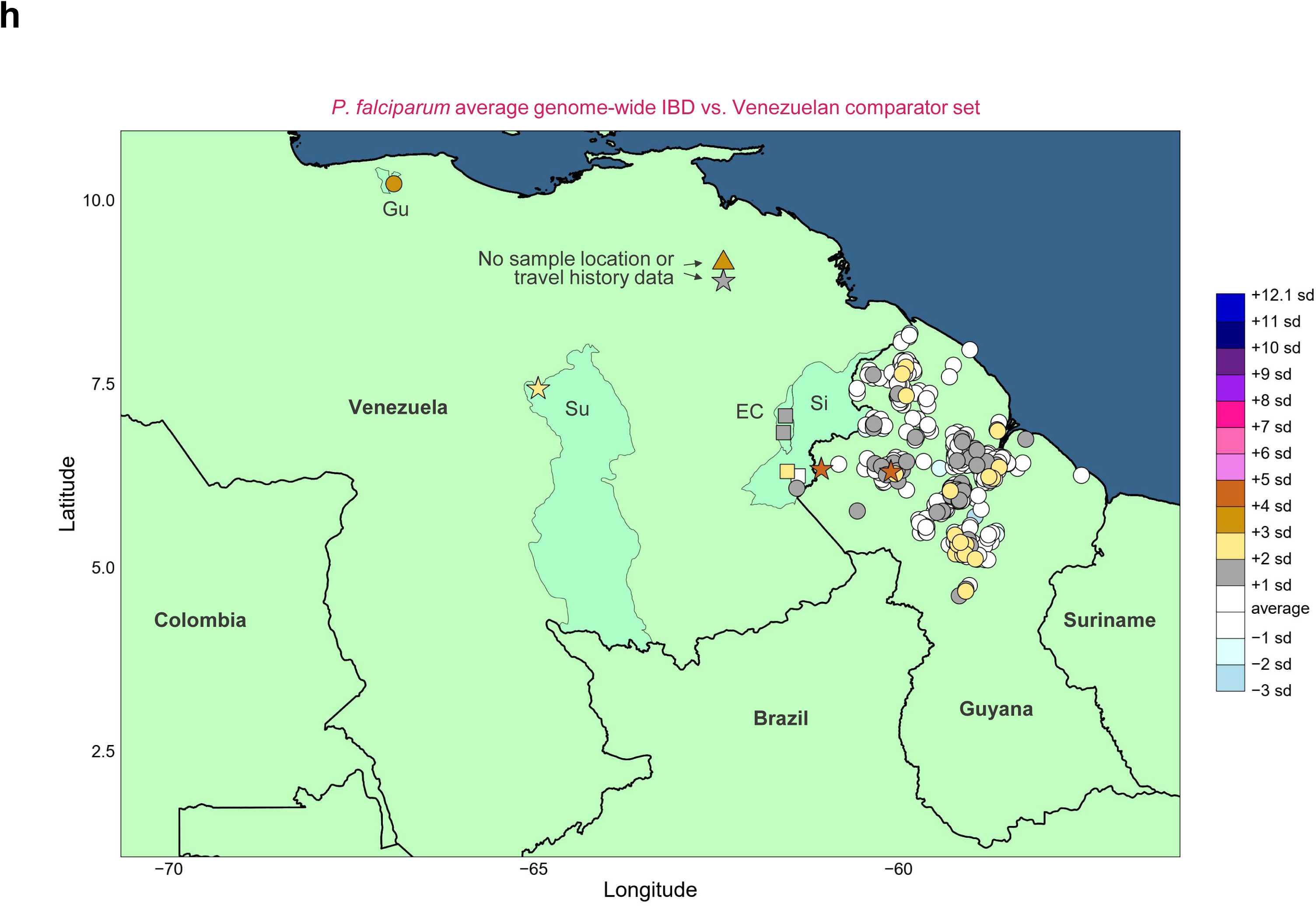
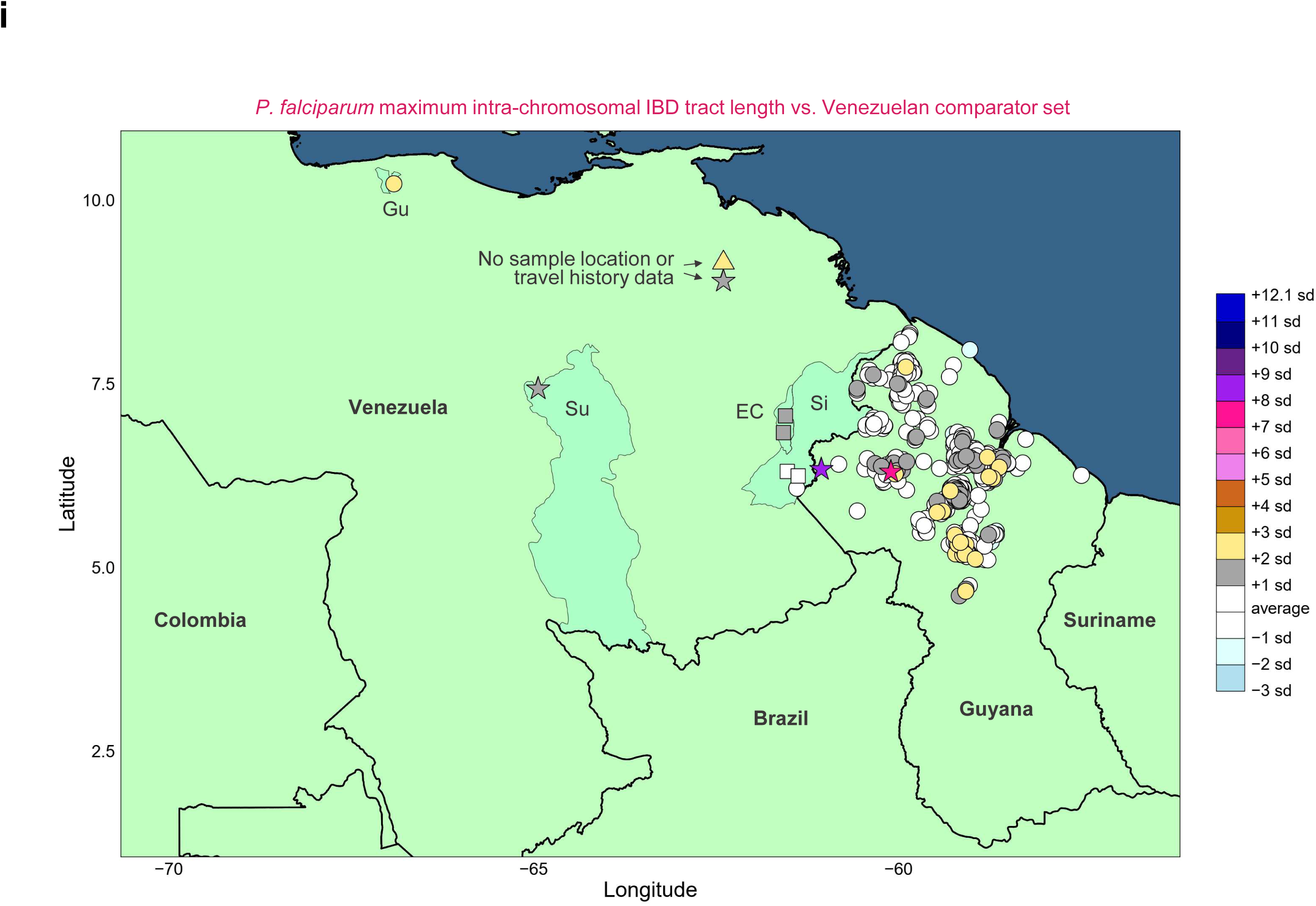
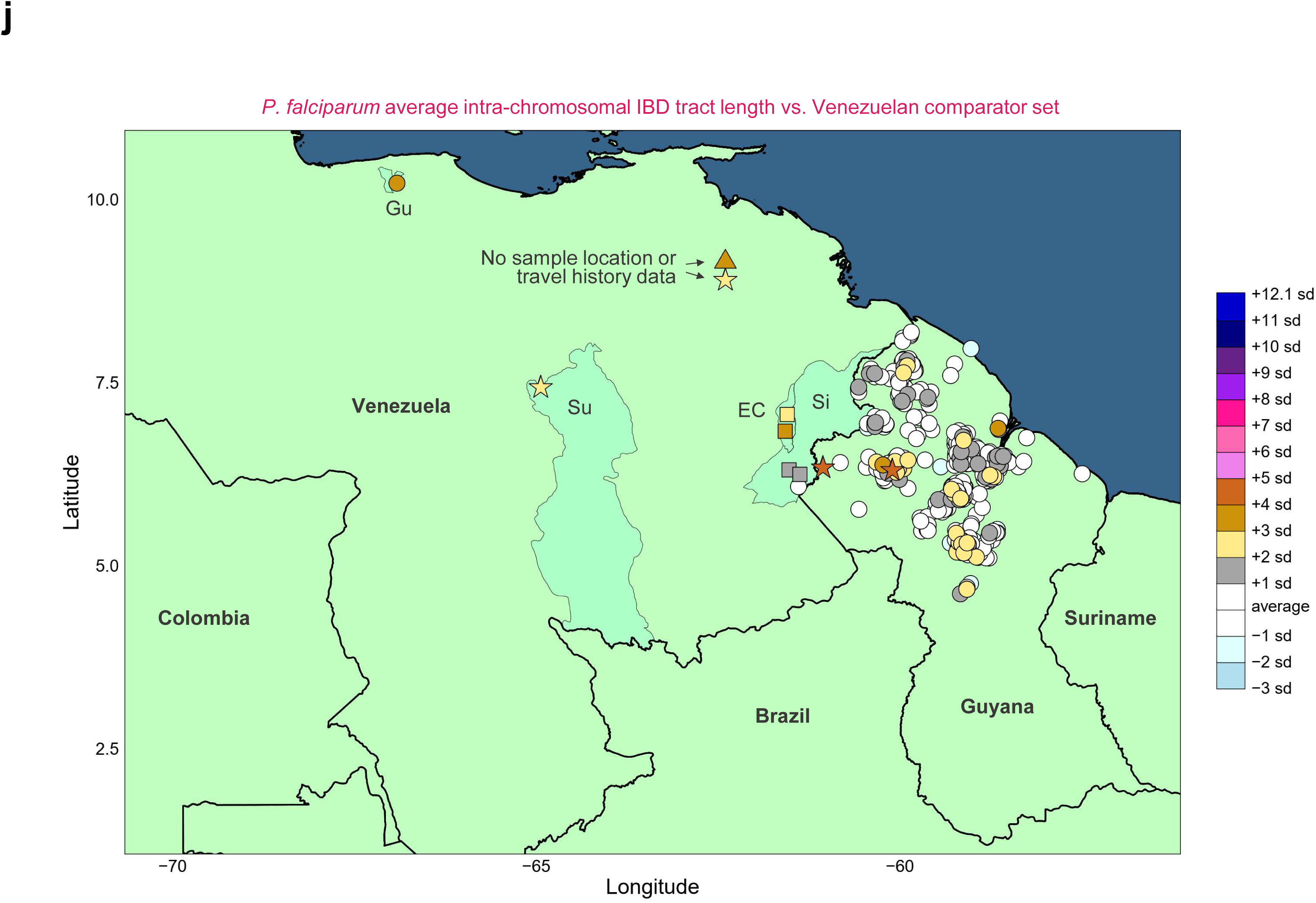
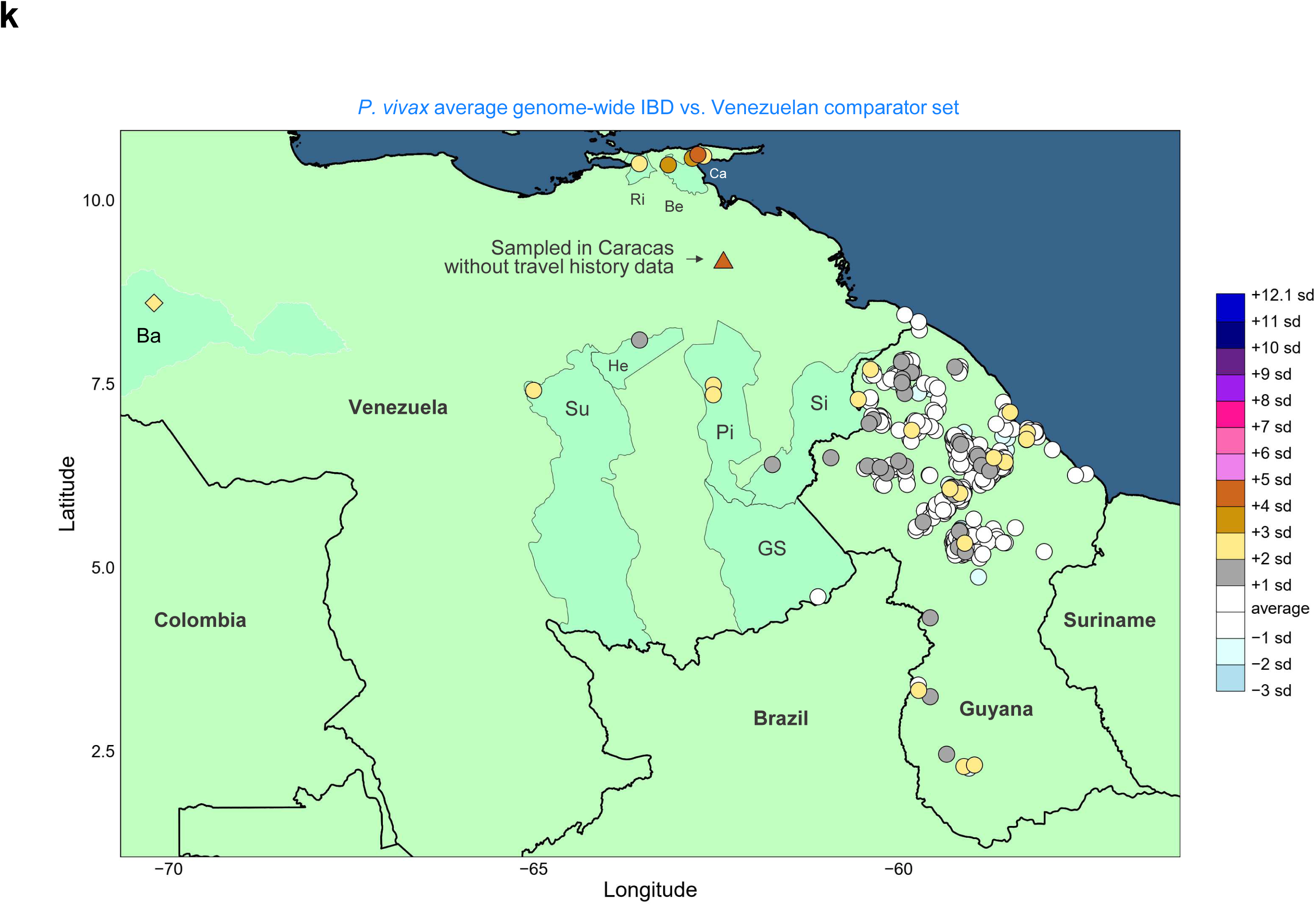
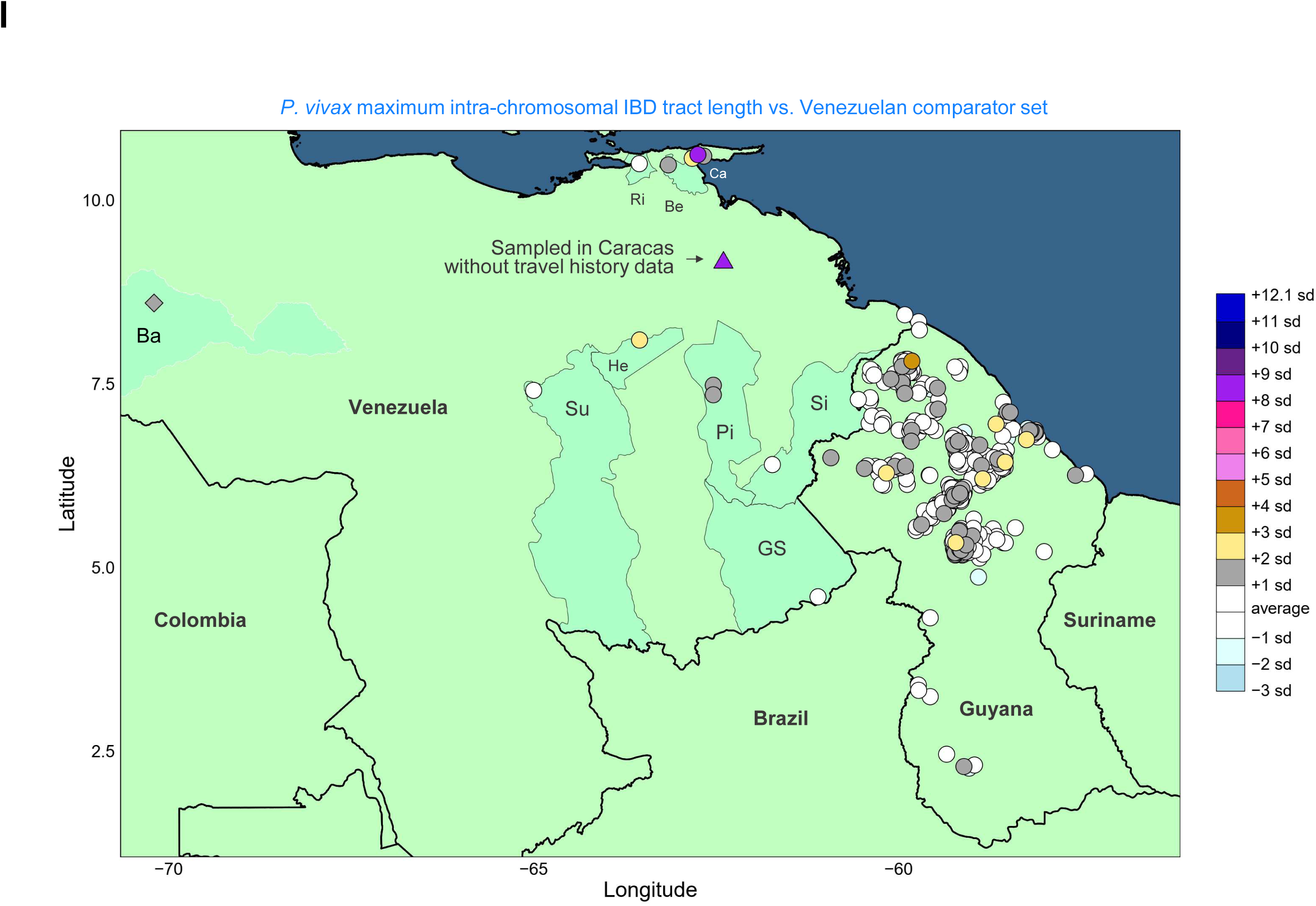
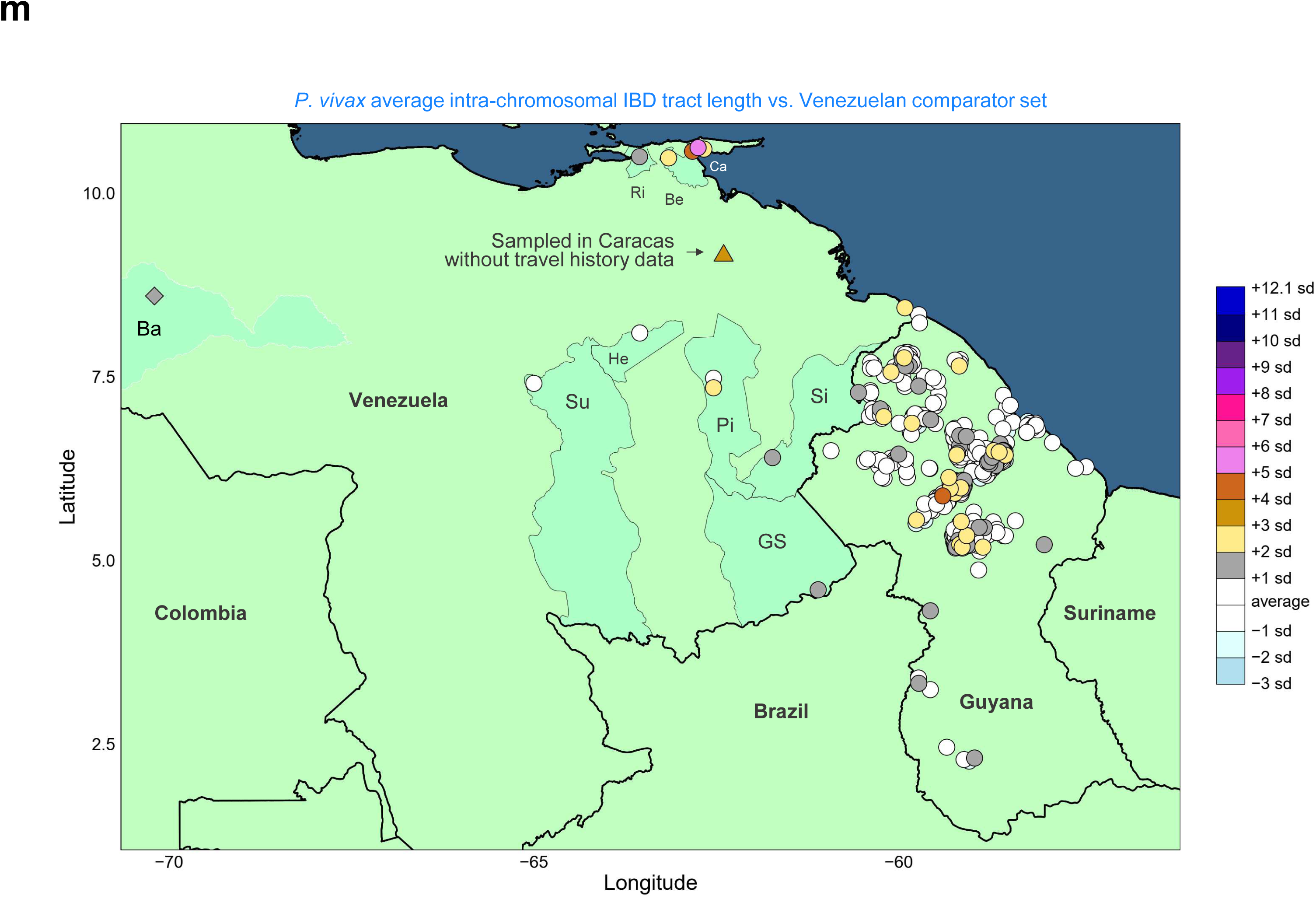
Differentiation between Guyanese and Venezuelan *P. falciparum* and *P. vivax* samples. Samples represent 2020-21 (Guyana) and 2015-16 + 2019 (Venezuela). **a)** IBD density curve for *P. falciparum* sample-pairs representing within-Guyana comparisons (G-G, solid line) and Guyana-Venezuela comparisons (G-V, dashed line). **b)** For *P. falciparum*, map of maximum genome-wide IBD vs. any Venezuelan sample (MaxVZ). **c)** For *P. falciparum*, map of genome-wide IBD vs. G4G410 (including comparison of G4G410 to itself (i.e., 100% IBD)). **d)** For *P. falciparum*, map of genome-wide IBD vs. G4G1043 (including comparison of G4G1043 to itself (i.e., 100% IBD)). **e)** IBD density curve for *P. vivax* sample-pairs representing within-Guyana comparisons (G-G, solid line) and Guyana-Venezuela comparisons (G-V, dashed line). **f)** For *P. vivax*, map of maximum genome-wide IBD vs. any Venezuelan sample (MaxVZ). **g)** Principal component analysis in *P. falciparum* (left) and *P. vivax* (right). Sample points plotted on PC1 vs. PC2 are colored according to epidemiological zone, and shape indicates country. All Venezuelan samples are classified to a single zone (yellow triangles). Analysis uses sites with >2% minor allele frequency and no missing genotype calls. **h)** For *P. falciparum*, map of average genome-wide IBD vs. any Venezuelan sample. **i)** For *P. falciparum*, map of maximum intra-chromosomal IBD tract length vs. any Venezuelan sample. **j)** For *P. falciparum*, map of average intra-chromosomal IBD tract length vs. any Venezuelan sample. **k)** For *P. vivax*, map of average genome-wide IBD vs. any Venezuelan sample. **l)** For *P. vivax*, map of maximum intra-chromosomal IBD tract length vs. any Venezuelan sample. **m)** For *P. vivax*, map of average intra-chromosomal IBD tract length vs. any Venezuelan sample. In each map, points are colored based on the standard deviation of the IBD metric from the mean (see color scale at right). Points with higher values are plotted above points with lower values, i.e., are not obscured. Circles represent samples with travel history recorded to the locality level (i.e., using specific coordinates). Squares represent samples with travel history recorded to the municipality level (coordinates are placed within known malaria areas of the municipality (black perimeter)). Travel history for sample CEM526_Pv-2 (diamond) is recorded only to the state level (Barinas (Ba), white perimeter)). Its coordinates are placed arbitrarily within Ba. Samples CEM541_Pv-9, PW0065-C, and SPT26229 (triangles and/or text annotation) do not contain any travel history details. Star symbols are used for clonal samples with representation in both Guyana and Venezuela. Municipalities are abbreviated within the Venezuelan states of Bolívar (EC (El Callao), GS (Gran Sabana), He (Heres), Pi (Piar), Sifontes (Sifontes), Su (Sucre)), Sucre (Be (Benítez), Ca (Cajigal), and Ri (Ribero)), and Miranda (Gu (Guaicaipuro)).

**Supplementary Fig. 10.**
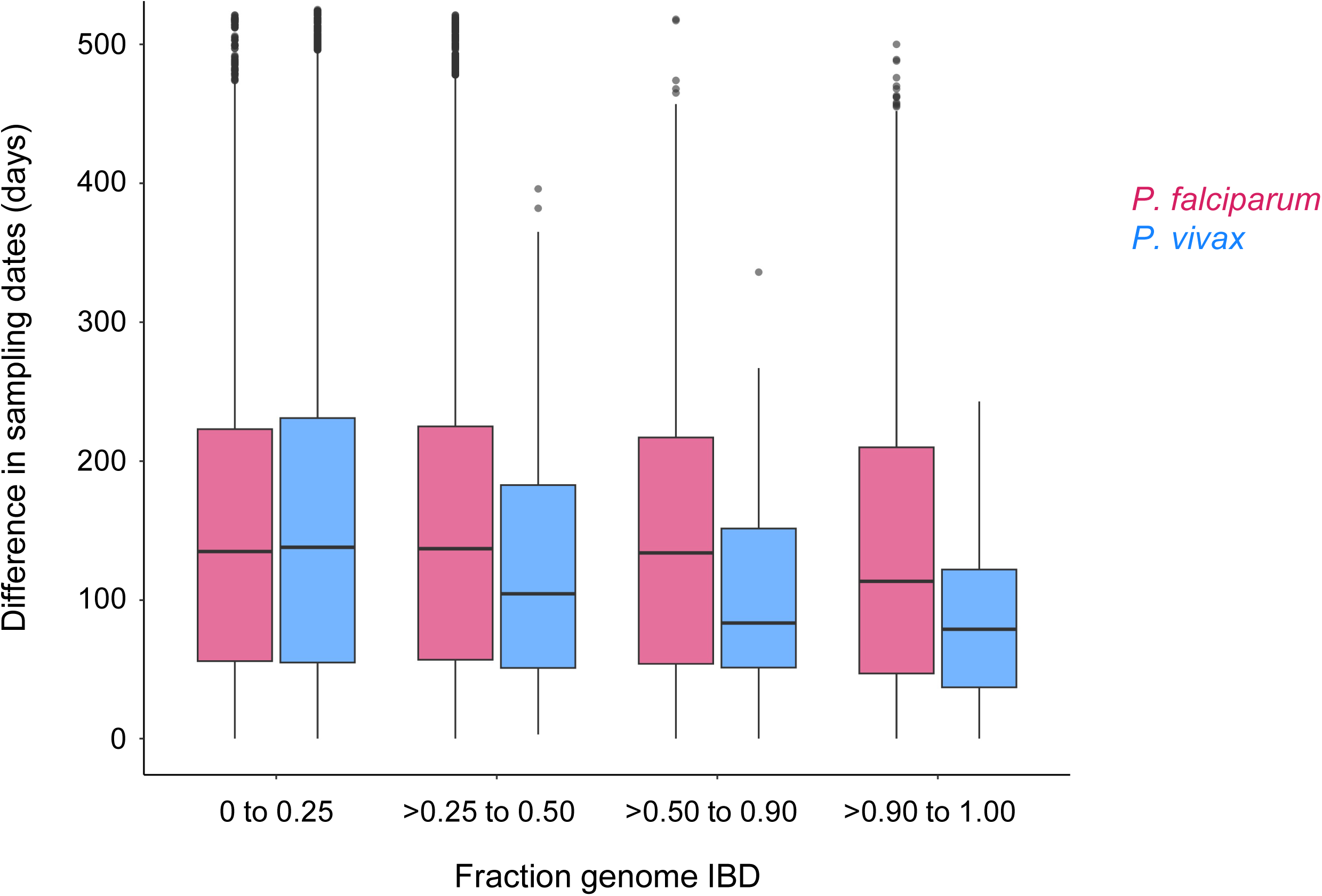
*P. falciparum* and *P. vivax* IBD with respect to time difference in sampling dates in Guyana. Boxplots summarize variation (median and quartiles) in time (days) between sampling for different IBD categories (increasing from low to high (clonal) on x-axis) in 2020-21.

**Supplementary Fig. 11.**
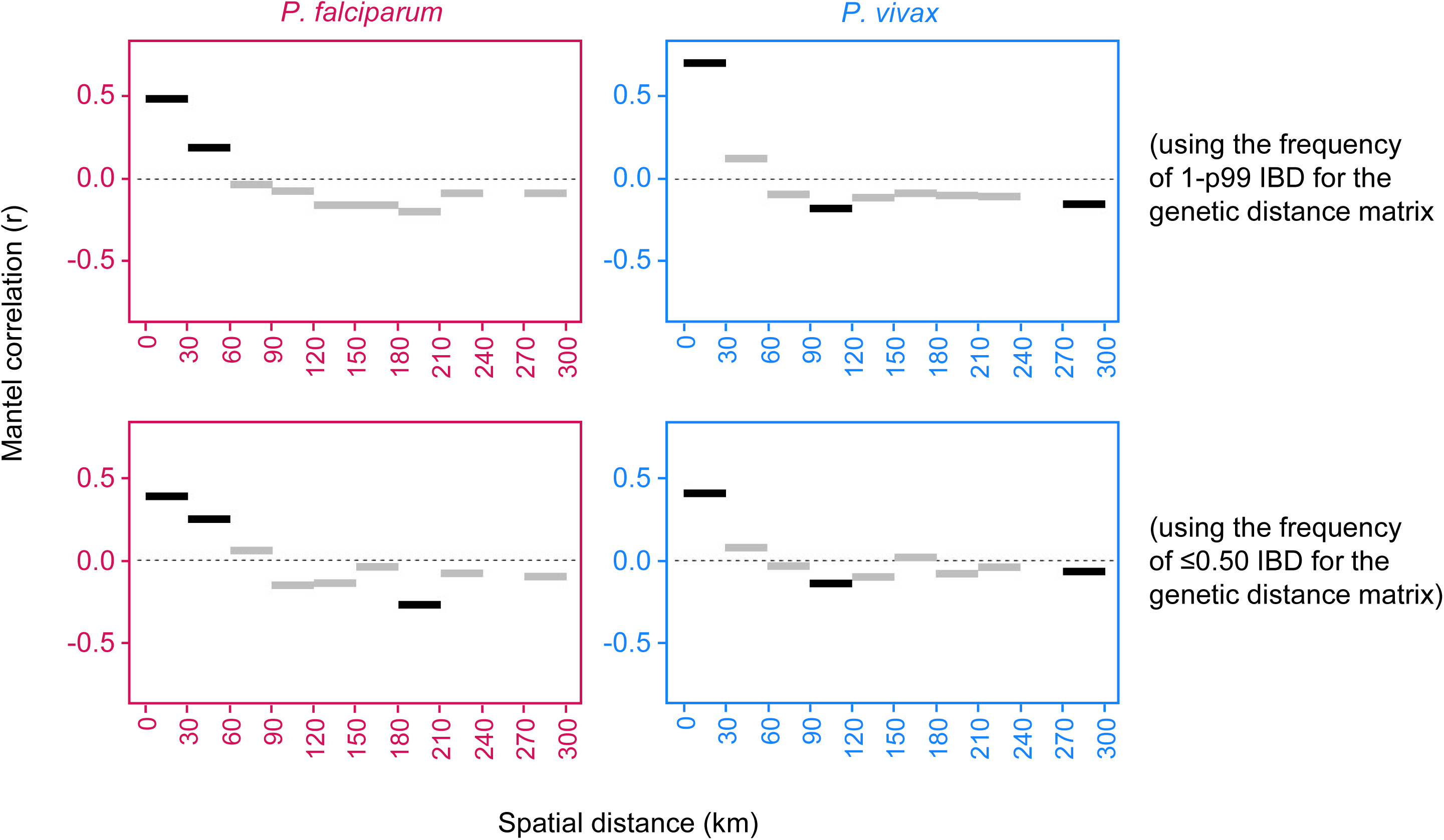
Mantel correlograms for *P. falciparum* and *P. vivax* in Guyana. Mantel correlation coefficients (r) are plotted on the y-axis for successive (non-overlapping) spatial distance classes (bars) separating infection localities in 2020-21. Black bars indicate significant r (p < 0.05) between genetic and spatial distance matrices. Top plots use 1-p99 IBD frequency and bottom plots use ≤0.50 IBD frequency for the genetic distance matrix. Positive r represents isolation-by-distance, i.e., that genetic distance increases with spatial distance.

**Supplementary Fig. 12.**
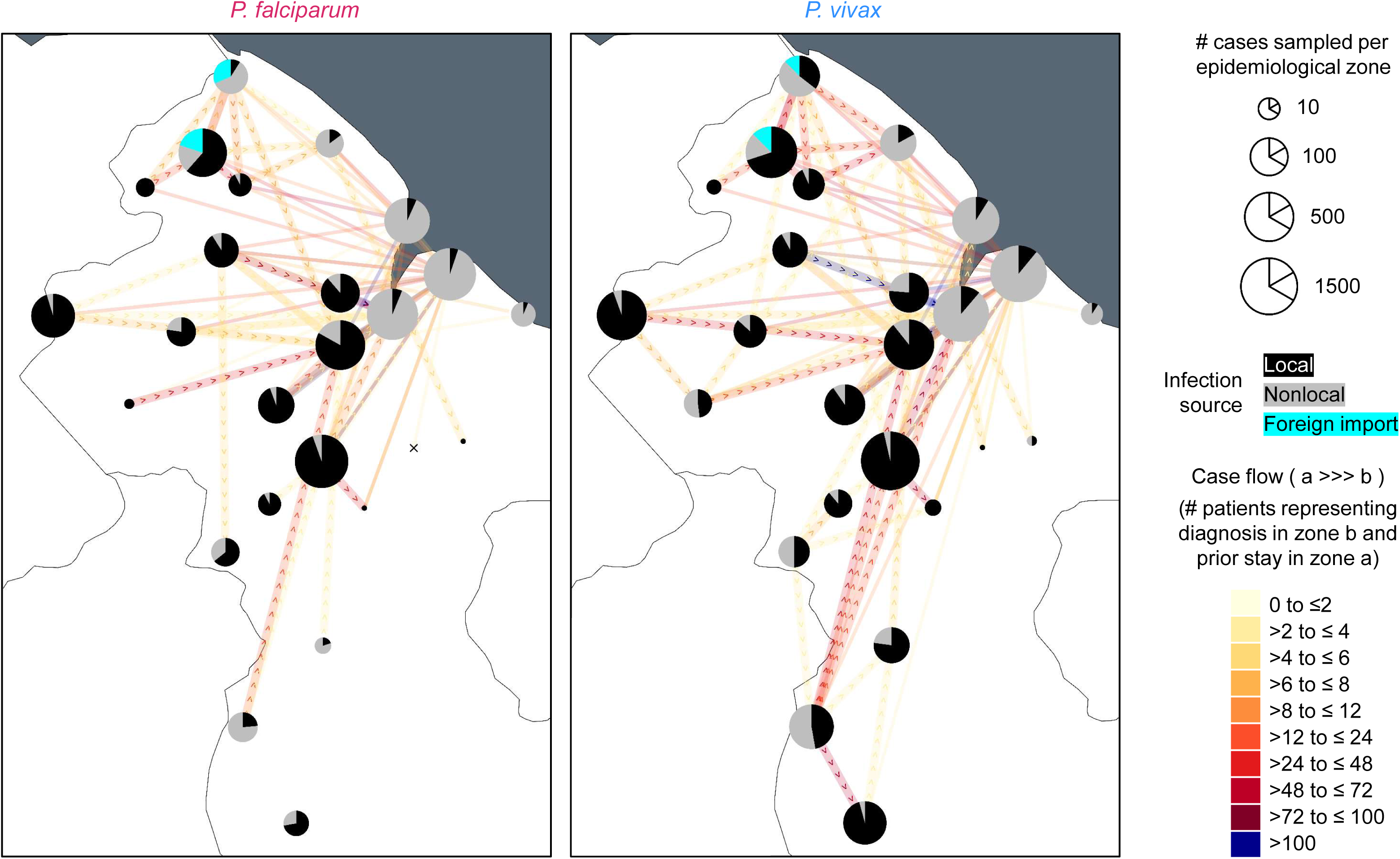
*P. falciparum* and *P. vivax* case flow in Guyana. Pie icon sizes represent the number of cases sampled within each epidemiological zone in 2019. Black slices represent the fraction of cases representing patients that reported having stayed in the same epidemiological zone 2 weeks prior to diagnosis. Grey slices represent the fraction of cases representing patients that reported having stayed in a different epidemiological zone 2 weeks prior to diagnosis. Case flow segments are mapped for such ‘non-local’ cases. These segments connect the reported location of prior stay with arrows to the location of diagnosis. Arrows are used if the location of diagnosis is considered an endemic zone (e.g., for this reason only solid lines connect to coastal zones around Georgetown). The number of case flow events recorded between zones is represented by segment color, increasing from yellow through orange and red to blue (see scale). The analysis uses 13,641 cases for which information on location of diagnosis and on location 2 weeks prior to diagnosis is available.

**Supplementary Fig. 13.**
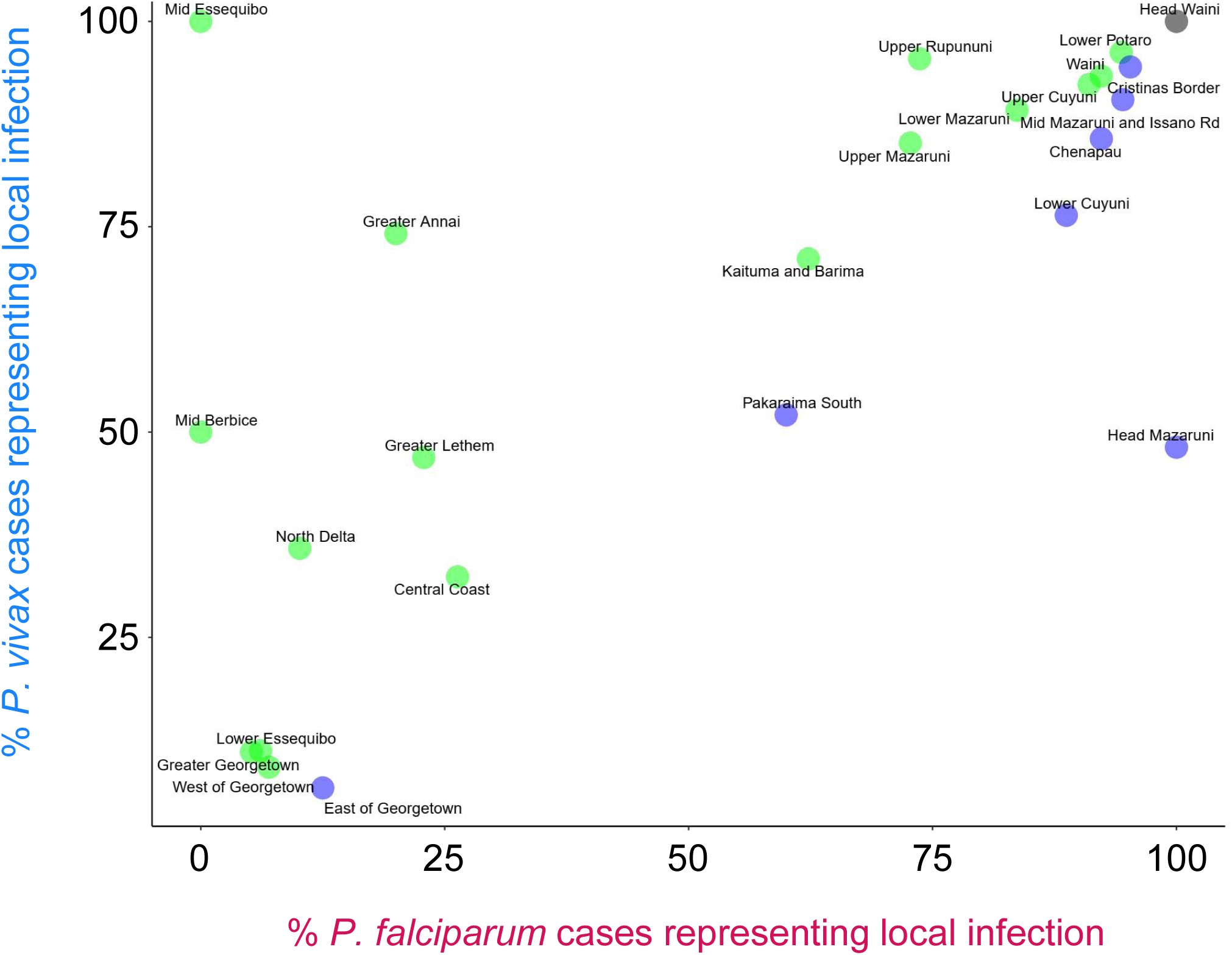
Rates of local infection for *P. falciparum* and *P. vivax* in Guyana. For each epidemiological zone (points), the percentage of *P. falciparum* cases representing local infection (x-axis) is plotted against the percentage of *P. vivax* cases representing local infection (y-axis) in 2019. Values correspond to black pie slices in Supplementary Fig. 12. Blue is used when higher values occur for *P. falciparum*, green is used when higher values occur for *P. vivax*, and black is used when values are equal between species.

**Supplementary Fig. 14.**
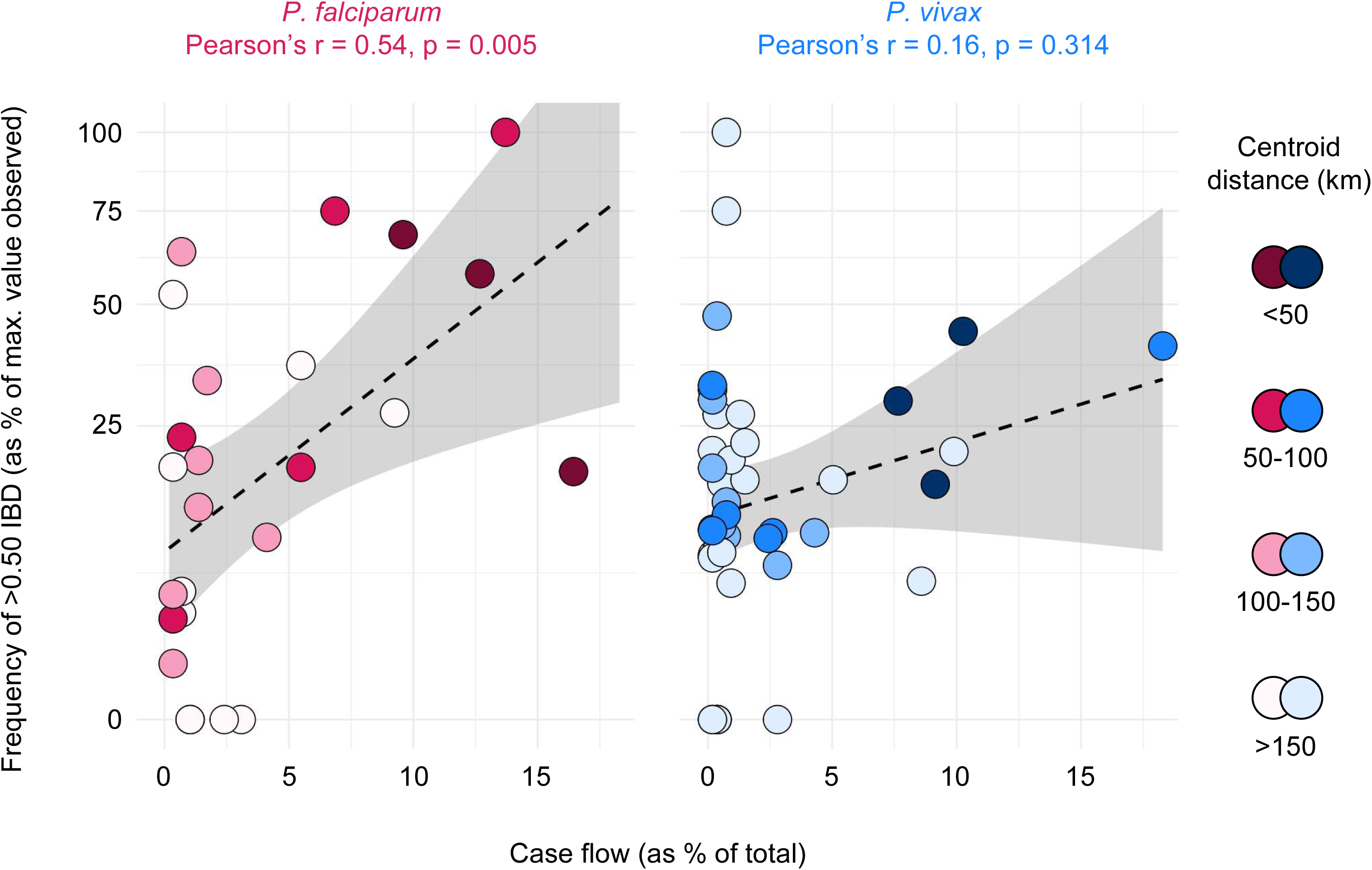
Relationship between the relative frequency of parasite p99 IBD and patient case flow detected between epidemiological zones in *P. falciparum* and *P. vivax* in Guyana. Genetic data (y-axis) represents 2020-21. Epidemiological (case flow) data (x-axis) represents 2019. Point color indicates the spatial distance separating the zones being compared. Comparisons represented by ≤50 comparisons are excluded. Pearson correlation is statistically significant in *P. falciparum* but not in *P. vivax*. Grey shading indicates 95% confidence intervals predicted by linear regression (dashed line).

**Supplementary Fig. 15.**
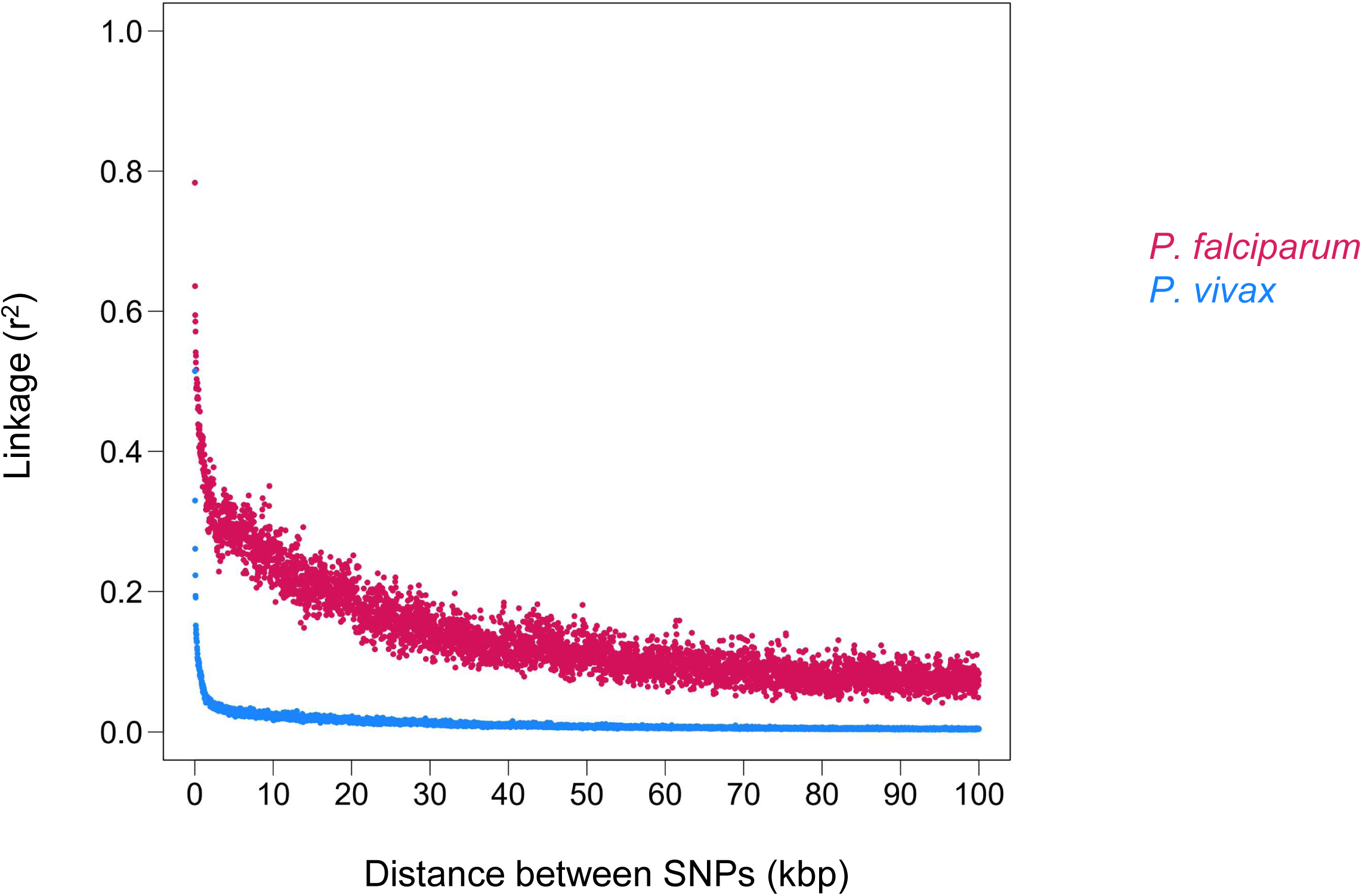
Linkage decay in *P. falciparum* and *P. vivax* in Guyana. Average linkage values (r^2^) between SNP sites are plotted in sliding 10 kbp windows (step size = 200 bp) for samples from 2020-21.

**Supplementary Fig. 16.**
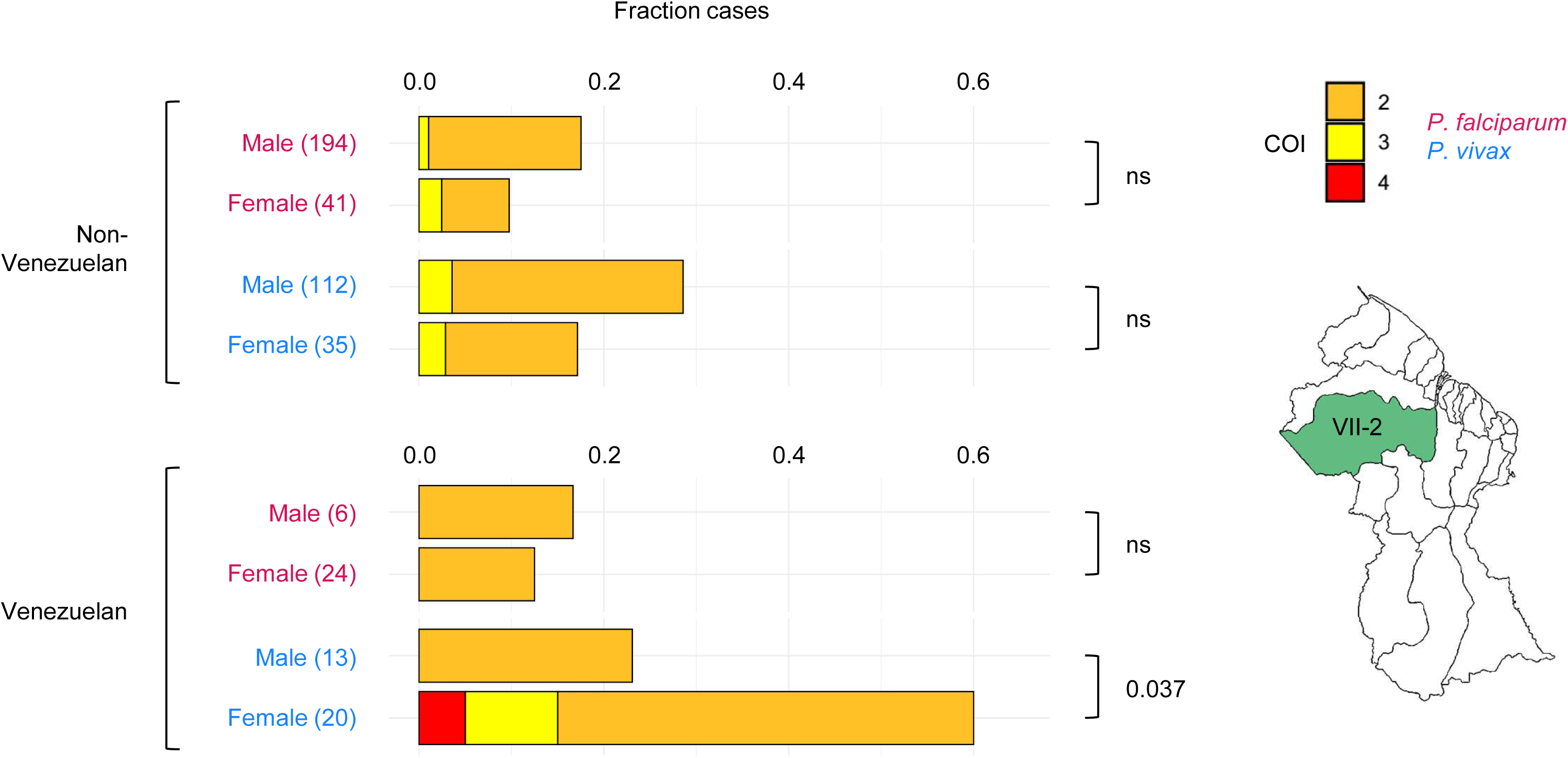
Complexity of infection values for *P. falciparum* and *P. vivax* in Venezuelan and non-Venezuelan patients with infections attributed to NDC VII-2 in Guyana. NDC VII-2 is highlighted in green in the map. Upper plots represent patients of non-Venezuelan nationality and lower plots represent patients of Venezuelan nationality (see large brackets at left). For each malaria species and patient gender, horizontally stacked bars represent complexity of infection (COI) values contributing to total observed polyclonality rate (full length of bar on x-axis) in 2020-21. Orange = 2 strains, yellow = 3 strains, and red = 4 strains. *P. vivax* polyclonality rate is significantly elevated in female Venezuelan patients vs. male Venezuelan patients (Chi-squared test). Differences are non-significant (ns) for other indicated comparisons (see small brackets at right).

**Supplementary Fig. 17.**
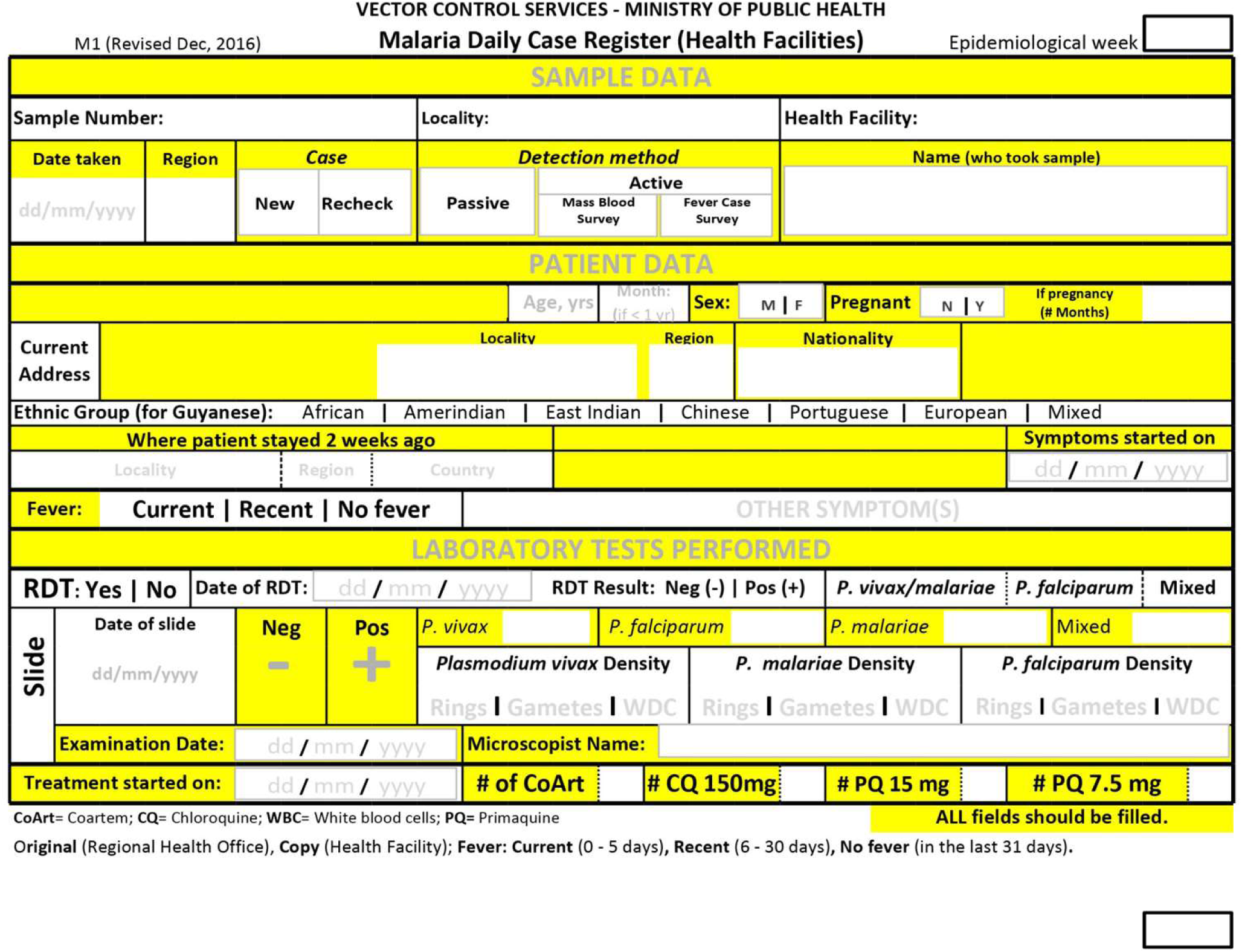
Malaria case form underlying patient metadata records analyzed from Guyana. Form records of sampling date, sampling location, patient gender, patient age, patient nationality, patient ethnicity, location of patient stay 2 weeks prior to diagnosis, and infecting species (microscopy or RDT result) were provided to study authors using anonymized patient codes. Only cases marked ‘New’ and ‘Passive’ were analyzed. The same form was used for 2019 and 2020-21 sample sets.

